# Analytical Validation of a Highly Accurate and Reliable Next-Generation Sequencing-Based Urine Assay

**DOI:** 10.1101/2024.04.05.24305286

**Authors:** Mara Couto-Rodriguez, David C Danko, Heather L Wells, Sol Rey, Xavier Jirau Serrano, John Papciak, P Ford Combs, Gabor Fidler, Christopher E. Mason, Caitlin Otto, Niamh B. O’Hara, Dorottya Nagy-Szakal

## Abstract

Culture is currently the gold standard for diagnosis of urinary tract infections (UTIs); however, it has poor sensitivity detecting urogenital pathogens, especially if patients have already initiated antimicrobial therapy, or have an infection from an organism that is not commonly cultured. False negative urine culture results can lead to the inappropriate use of antimicrobial therapies or to the progression to urosepsis in high-risk patients. Though not commonly applied to urine in a clinical setting, Next-generation sequencing (NGS)-based metagenomics offer a solution as a precision diagnostic. We developed and validated BIOTIA-ID, a clinical-grade NGS-based diagnostic pipeline for the detection and identification of pathogens in urine specimens. Remnant clinical urine specimens, and contrived sterile urine spiked with common UTI pathogens, were processed with our end-to-end assay including extraction, metagenomic library preparation and Illumina NextSeq 550 sequencing. We trained and applied a bioinformatic pipeline that uses machine learning (ML) to identify pathogens. Internal controls and other quality control measures were incorporated into the process to provide rigorous and standardized results. The assay was tested on 1,470 urine specimens and achieved 99.92% sensitivity, 99.95% specificity and a limit of detection (LoD) of <25,000 CFU/mL and <5,000 CFU/mL in bacteria and fungi, respectively. Discordant results were reconciled with additional testing by target-specific qPCR or 16S Sanger sequencing; 87% of the NGS results were ultimately determined to be the correct result. Overall, these data demonstrate that BIOTIA-ID is a highly accurate clinical-grade diagnostic tool with notable advantages over current culture-based diagnostics.

**Conflict of Interest Statement:** MCR, DCD, HLW, SR, XJS, JP, PFC, GF, CEM, CO, NBO and DNS are employees at Biotia, Inc.

## INTRODUCTION

Annually, 11 million people in the United States and 404 million people worldwide are diagnosed with a urinary tract infection (UTI) (1, 2). Immunocompromised patients and patients with urological conditions are at higher risk to develop complicated and/or recurrent UTI (cUTI/rUTI) as well as to progress to urosepsis, resulting in higher morbidity and mortality and increased cost of care(3–8). In hospitalized patients, UTIs are associated with 2.3% of the mortality rate and an estimated annual cost of $340 to $450 million in the United States alone(1). Approximately 30% of septic patients have an infection originating from the urogenital tract, with a multinational study revealing that 12% of nosocomial UTI patients progressed to urosepsis (7, 9). Rapidly identifying UTI pathogens in high-risk patients is crucial to administering an appropriate treatment and minimizing unnecessary broad-spectrum antibiotic use.

Urine culture is the standard of care (SOC) for the detection and identification of pathogens causing UTIs. However, culture-dependent approaches are focused on an Enterobacterales-centric paradigm which is based on assumptions that urine is sterile (10–14), disregards mixed infections as contamination (15–17), and is based on clinical studies employing heterogeneous definitions and thresholds for a UTI (18, 19). Culture has several limitations, including: (1) an inability to identify hard-to-grow microbial organisms, such as anaerobes and fungi; (2) challenges in identifying rare pathogens or organisms not routinely cultured; and (3) risk of false negative results for patients being treated with antibiotics (20–23). Long diagnostic turnaround time (TAT) or inconclusive results could lead to the empiric treatment of suspected infections with broad-spectrum antibiotics that are often inappropriate and may contribute to increased rates of drug resistance (24, 25).

The emergence of new molecular and culture-based techniques has underscored the drawbacks of traditional culture and challenged our understanding of UTIs, raising questions around what is considered a urogenital pathogen and the role of mixed bacterial communities in UTIs(12, 20, 26–32). Genomics-based assays enable direct specimen analysis without the need for culturing by sequencing genetic material, which is analyzed for the presence of pathogenic organisms through detection of their DNA. By comparing detected genetic sequences to a comprehensive microbial genome database, genomics-based tests can accurately and rapidly identify pathogens in cUTI samples, which may have atypical pathogens, multiple co-infecting organisms, and complex drug resistance profiles. Concerns have been raised about the sensitivity of molecular approaches to differentiate between opportunistic and commensal pathogen detection and differentiation of colonization versus infection (30, 33–36). However, careful development of genomics-based assays that center on extensive validation leveraging culture, molecular and bioinformatics techniques can increase the utility of this methodology in routine clinical practice, thus offering promising supplemental testing method to the current gold standard UTI testing.

Here, we developed, optimized, and clinically validated a (NGS)-based urine assay, BIOTIA-ID, that has high diagnostic sensitivity and specificity and provides comprehensive detection of urogenital pathogens within a single test.

## MATERIALS AND METHODS

### Clinical Validation Strategy

All specimens were processed with our version-controlled BIOTIA-ID Urine NGS Assay laboratory protocol, analytical pipeline, and reference microbial database in Biotia’s CLIA-certified laboratory. The assay and clinical validation were designed based on New York State Department of Health validation guidelines for submission of a Laboratory Developed Test for bacteriology and mycology nucleic acid amplification assays. An overview of the assay workflow and clinical validation design is illustrated in **Figure 1**.

**Figure 1.**
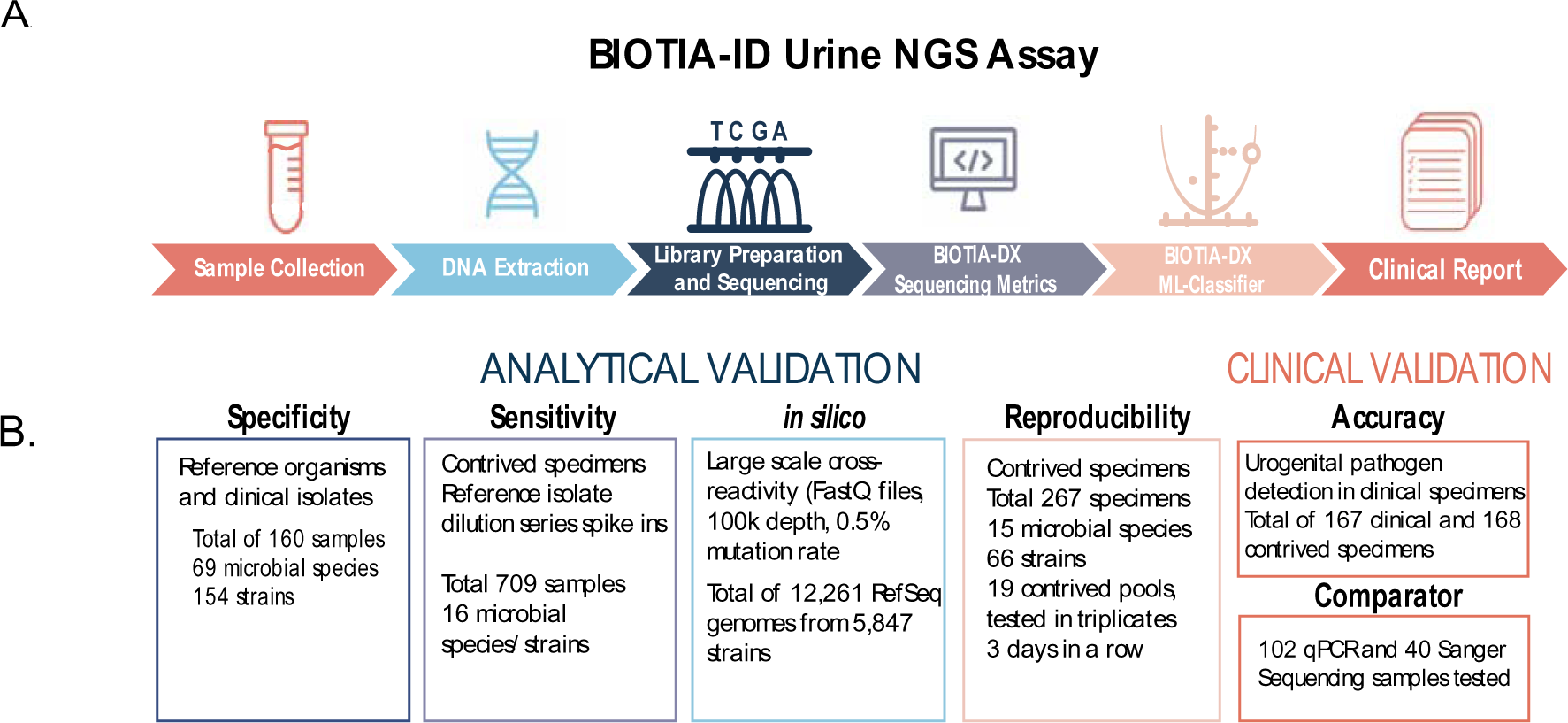
**(A)** An overview of the BIOTIA-ID Urine NGS Assay workflow, consisting of sample collection (urine specimens in standard UTT), nucleic acid extraction, library preparation and sequencing using the Illumina platform, BIOTIA-DX metagenomic analysis, multiple decision tree-based machine learning modeling and generation of a clinical report with urogenital pathogens detected. **(B)** Analytical validation including specificity, sensitivity and *in silico* studies evaluated the clinical performance characteristics of the assay. The clinical validation established the accuracy of urogenital pathogen detection in clinical specimens with additional comparator studies using qPCR and Sanger sequencing in cases where culture and NGS results disagreed. Furthermore, we completed an inter/intra assay reproducibility study to show the validity and consistency of our results.

### Clinical Specimens and Reference Materials

Deidentified urine specimens in urine transport tubes (UTT) were collected as residual samples after routine clinical testing from different microbiology reference laboratories and were provided with the culture results for each specimen following their standard operating procedures to report pathogens. Samples were collected and processed under an institutional review board (Advarra Pro00038083) without demographic information. A collection of clinical and reference isolates (ATCC, Zeptometrix) was also used for the clinical validation including bacteria, fungi, viruses, and parasites (listed in **Table 1**). Reference and clinical microbial isolates used in the contrived specimens were inoculated and grown in Tryptic Soy Agar with 5% Sheep Blood at 37°C for 24-48 hours.

**Table 1.** Reference organisms and clinical isolates tested and validated on the specificity study. Whole organisms of urogenital pathogens detected by the assay, related organisms, and other organisms that can be present in urine specimens were tested by spiking microbial cells into a negative urine matrix (50,000 CFU/mL for bacteria and 25,000 CFU/mL for fungi). A total of 69 microbial species (bacteria, fungi, viruses and parasites) and 154 strains were evaluated. (*) Indicates organisms tested with BIOTIA-ID assay but not reported as a key urogenital pathogen.

| Gram Negative Enterobacterales | Gram Negative non Enterobacterales | Gram Positive | Fungi |
| --- | --- | --- | --- |
| *Citrobacter koseri* | *Acinetobacter calcoeticus-baumannii-complex* | Other *Staphylococcus* spp. | *Candida albicans* |
| *Citrobacter freundii* | *Acinetobacter lwoffii* | Anginosus group *Streptococci* | *Candida auris* |
| *Enterobacter cloacae-complex* | *Pseudomonas aeruginosa* | *Streptococcus agalactiae* | *Candida dubliniensis* |
| *Escherichia coli* | *Strenotrophomonas maltophilia* | *Streptococcus mitis* | *Candida glabrata* |
| *Klebsiella aerogenes* | **Anaerobic bacteria** | **Other Bacteria** | *Candida guilliermondii* |
| *Klebsiella oxytoca* | *Anaerococcus* spp. | *Chlamydia trachomatis* * | *Candida kefyr* |
| *Klebsiella pneumoniae-complex* | *Bacteroides fragilis* | *Gardnerella vaginalis* | *Candida krusei* |
| *Klebsiella variicola* | *Prevotella* spp. | *Mycoplasma genitalium* * | *Candida lusitanae* |
| *Morganella morganii* | **Gram Positive** | *Mycoplasma hominis* * | *Candida parapsilosis* |
| *Proteus mirabilis* | *Aerococcus* spp. | *Neisseria gonorrhoeae* * | *Candida tropicalis* |
| *Proteus vulgaris* | *Corynebacterium urealyticum* | *Treponema pallidum*.* | *Cryptococcus neoformans* * |
| *Providencia rettgeri* | *Enterococcus faecalis* | *Ureaplasma* spp.* | *Rhodoturula mucilaginosa* * |
| *Providencia stuartii* | *Enterococcus faecium* | **Viruses** | *Saccharomyces cerevisiae* * |
| *Raoultella ornithinolytica* | *Staphylococcus aureus* | CMV* | **Parasites\*** |
| *Salmonella Typhimurium* * | *Staphylococcus epidermidis* | HPV* | *Cryptosporidium* spp.* |
| *Serratia marcescens* | *Staphylococcus lugdunensis* | HSV1* | *Entamoeba* spp.* |
| *Shigella flexneri* * | *Staphylococcus saprophyticus* | HSV2* | *Giardia lamblia* * |
|  |  |  | *Trichomonas vaginalis* * |

### Clinical-Grade Metagenomic Sequencing for Infectious Disease Diagnostics

*BIOTIA-ID Urine NGS Assay.* Genomic DNA (gDNA) extraction from 2 mL of urine was performed with QIACube-MDx and yields were quantified with the Qubit Flex. Normalized gDNA was spiked with 5% internal positive control (IPC) and processed with the Illumina DNA prep library preparation kit. Libraries were quality checked for size and concentration with Tapestation 4200 and Qubit Flex, respectively. Libraries were pooled in equimolar concentrations and sequenced on an Illumina NextSeq 550 platform.

*Assay Controls.* Each NGS run includes three external controls and one internal control, which are processed with each batch of samples. The external controls consist of a no template control (NTC, nuclease free water), positive control (PC, Zymobiomics) and negative extraction control (NEC, negative urine matrix). The IPC is a proprietary gDNA that is spiked into each sample at 5%.

*BIOTIA-DX Software.* After sequencing, the FastQ files were first subjected to a filtering step which removed low-quality reads and sequencing adapters. Detection of the internal positive control was performed for each sample as a confirmation of overall sequencing quality. Human sequence data was then removed from downstream analysis. The remaining reads were first compared against a large database of microbial genomes in a coarse classification step using a commonly used metagenomics tool, *kraken2* (37). We use a custom *kraken2* microbial database within BIOTIA-DX that includes curated, high-quality genetic sequences from bacterial, fungal, viral, and non-fungal eukaryotic parasitic organisms. In total, more than 7,000 organisms are represented in the database, with diverse representation of microbial species and strains.

Despite any degree of database curation, metagenomics tools often result in spurious, false positive hits, which has historically made it difficult to use NGS in a clinical diagnostic setting(38–41). To distinguish between clinically relevant infections, off-target spurious hits by the metagenomics software, or low levels of cross-contamination, we designed a classifier using ML that estimates the probability that a microbial organism is present in a sample. Targets exceeding predefined *kraken2* thresholds in the coarse classification step were tested using this ML classifier in a fine classification step. Finally, organisms which exceeded a pre-defined probability threshold from the classifier were called as present in the sample.

### Pre-Validation (BIOTIA-DX Training Set)

To ensure clinical-grade diagnostic accuracy of these detection thresholds, we trained our ML classifier to recognize clinical-level infections using a training set of clinical samples with known and orthogonally validated infectious organisms. We sequenced and tested a pre-validation sample set of 114 clinical urine samples with known organisms identified by culture, and in cases where our algorithm results differed from culture results, we used PCR testing as a comparator assay. Results from PCR tests were then used to update the list of known organisms in the pre-validation set and re-train the ML classifier with the updated results. The classifier uses a variety of bioinformatic features chosen specifically to distinguish true infections from false positives. These features were calculated for each organism being tested and included statistics such as the depth and evenness of genome coverage, read identity to reference genomes, and the quality of assembled contigs.

### Analytical Validation

*Specificity.* Reference organisms and clinical isolates were tested and validated to establish the specificity of the assay. Whole organisms of urogenital pathogens detected by the assay, genetically related organisms, and other organisms that can be present in urine specimens were tested by spiking microbial cells into negative urine matrix. A total of 69 microbial species (bacteria, fungi, viruses, and parasites) and 154 strains were evaluated (**Table 1**). Microbial isolates were spiked into a negative urine matrix at concentrations of 25,000-50,000 CFU/mL and processed with the assay.

*Analytical Sensitivity.* The analytical sensitivity was assessed in 16 of the most prevalent urogenital pathogens, by spiking reference whole organisms into negative urine matrix (**Bacteria**: *Acinetobacter baumanii*, *Bacteroides fragilis*, *Escherichia coli, Enterococcus faecalis, Gardnerella vaginalis, Klebsiella pneumoniae, Prevotella spp., Proteus mirabilis*, *Pseudomonas aeruginosa* and *Staphylococcus aureus;* **Fungi:** *Candida albicans, Candida auris, Candida glabrata, Candida krusei, Candida parapsilosis, Candida tropicalis*). Serial dilutions of each pathogen were spiked in triplicate followed by gDNA extraction. The limit of detection was defined when the target organism is detected in ≥95% of the samples.

*In silico Analytical Specificity.* To assess the analytical specificity of our pipeline, we used a total of 12,264 RefSeq genomes from 5,847 unique bacteria, eukaryotic organisms, and viruses, to simulate sequencing files with 100,000 reads each. A random mutation rate of 0.5% was applied during simulation to introduce a reasonable degree of variability. Simulated read files were then run through BIOTIA-DX in an identical manner to sequenced laboratory samples and tested for key organisms to assess the potential for organisms outside our laboratory sample set to generate false positive results for key organisms.

*Inter/Intra-Reproducibility.* For assessment of the reproducibility in bacterial detection, five different pools containing one clinical strain each of *E. coli, E. faecalis, K. pneumoniae, P. mirabilis* and *S. aureus* were tested. Each pool had different strains for each species (altogether 25 different strains) and were spiked into a negative urine matrix at a concentration of 25,000 CFU/mL per analyte. Each pool was then tested daily for three consecutive days in three technical replicates for a total of 15 replicates per pool), 45 replicates per analyte and 6 negative samples. A smaller follow up study sought to evaluate reproducibility of bacterial detection at different concentrations 100,000 CFU/mL (high), 25,000 and 20,000 CFU/mL (medium) and 15,000 CFU/mL (low). Two bacterial pools per concentration were tested in triplicate across three different days for a total of 9 replicates per concentration per analyte. A similar experimental design was performed to assess fungal reproducibility of ten fungal analytes. Five different pools each containing one different strain of *C. albicans*, *C. auris*, *C. glabrata*, *C. krusei*, *C. parapsilosis*, and *C. tropicalis* and a sixth pool with one strain of *C. dubliniensis, C. guilliermondii, C. kefyr* and *C. lusitaniae* were tested in triplicate across three different days. A total of 15 replicates per pool, 45 replicates per analyte and 27 negative samples were evaluated.

### Clinical validation

*Accuracy.* We tested a combination of urine clinical specimens and contrived samples (whole organism microbial reference strains and clinical isolates spiked into negative clinical matrix) to assess accuracy. Deidentified, remnant clinical urine samples were obtained from a reference laboratory and stored at -80°C until processing with the assay. Urine clinical samples were collected based on the pathogens diagnosed by culture. At least 30 specimens each with the most common urogenital pathogens (*E. coli, E. faecalis, K. pneumoniae, P. mirabilis* and *S. aureus*) were collected. When fewer than 30 samples for a given pathogen were available, contrived specimens were used to bring the total number of samples for that target to 30 (a total of 335 samples; 167 clinical specimens and 168 contrived). Contrived samples were used for *S. aureus* (n=30), *P. mirabilis* (n=9), *K. pneumoniae* (n=8), *C. albicans* (n=33), *C. auris* (n=30), *C. glabrata* (n=30), *C. krusei* (n=33), *C. parapsilosis* (n=34), *C. tropicalis* (n=30) with a spike in concentration close to the LoD (∼2.5 - 5× LoD). In addition, 35 specimens negative by culture were tested.

*Comparator Testing.* gDNA from urine specimens was used to perform confirmatory testing of clinical samples yielding results in which NGS and culture findings did not agree. Discrepant results were adjudicated either with qPCR or Sanger Sequencing. Taxa- or group-specific molecular assays were used to amplify analytes of interest (primers and references are listed in **Supplemental Table 1)**. PCR amplicons were sent for Sanger sequencing and consensus sequences were used to determine percent identity of the organism detected in the clinical specimen. A total of 8 different target-specific qPCR assays and 8 Sanger sequencing assays were used to test 123 discrepant taxa and 19 concordant taxa.

### Code Availability

The BIOTIA-DX software used is described in the Methods section. The software leverages the following openly available tools: fastp v0.23.3, bowtie2 v2.5.1, minimap2 v2.22, SNAP aligner 2.0.3, samtools v1.6, and kraken2 v2.1.3. The ML classifier is a proprietary portion of the code(37, 42–46).

### Data Availability

The data supporting the study findings are available from the corresponding author on request. Sequencing data that support the finding of this study (with human reads removed) have been deposited in GeoSeeq, a publicly available metagenomic platform, and available upon request.

## RESULTS

We developed an end-to-end urine clinical diagnostic metagenomics assay to detect urogenital pathogens and validated its performance on 1,470 samples sequenced in 65 sequencing runs (**Figure 1**). Clinical and analytical validation experiments were conducted with clinical and contrived specimens to assess specificity, sensitivity, BIOTIA-DX *in silico* performance, reproducibility, and accuracy (**Figure 1B**).

Microbial read counts, IPC detection, and external positive (PC) and negative controls (NEC and NTC) sequencing performance was assessed. Validation samples averaged 7.9M raw reads and 4.7M microbial reads after human removal (**Figure 2A**). The PC generated similar read depths as testing samples, while the NEC and NTC had reads < 750K. The IPC was detected within expected range (1-5% relative abundance), except in the negative controls (75% relative abundance) (**Figure 2B**).

**Figure 2.**
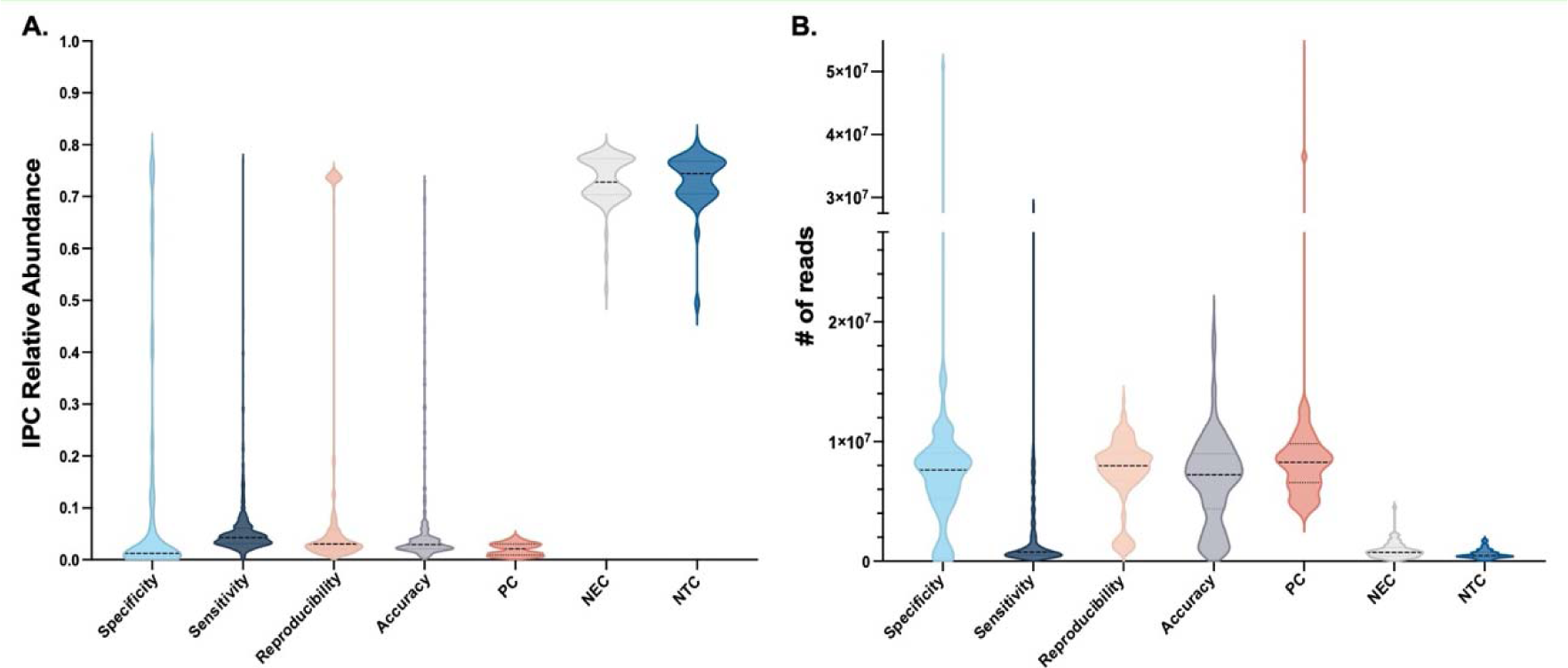
The assay performance of specimens processed in the analytical (specificity, sensitivity, and reproducibility) and clinical validation (accuracy) studies was defined by evaluating the internal positive control (IPC) and the number of microbial reads. **(A)** The IPC was detected in all specimens and positive controls (PC) at an expected range (1-5% abundance). The IPC reads represented the majority of the reads detected in negative extraction controls (NEC) and negative template controls (NTC). **(B)** Violin plots depicting the number of microbial reads obtained in each study and controls. As expected, the clinical and contrived specimens processed through the studies and the PCs generated similar microbial read depth (average 7.9M of microbial reads), while the NEC and NTC had microbial reads below 750k. Violin plots show IPC abundance and microbial read distribution for all samples and controls processed in each study: Specificity (light blue, n=160); Sensitivity (navy blue, n=709); Reproducibility (light pink, n= 267); Accuracy (gray, n= 325), PC (dark pink, n=65), NEC (light gray, n=130), NTC (cerulean blue, n=130)

### Key performance characteristics of BIOTIA-ID. SPECIFICITY

The similarity among different microbial genomes, the diversity of clinical isolates from reference genomes and contaminating DNA fragments impacts the analytical sensitivity of a metagenomic test. Urogenital pathogens detected by the assay and genetically related organisms were processed with BIOTIA-ID (**Table 1**), yielding a sensitivity and specificity of 100% and 99.96% for bacterial targets and 100% sensitivity and specificity for fungal targets. During assay development, several taxa not extensively represented in the clinical training set generated false positive and false negative results leading us to process 64 additional species in 80 contrived urine samples to feed more training data to the model. This approach coupled with hyperparameter tuning of the model ensued the increased sensitivity and specificity reported above.

## SENSITIVITY

The analytical sensitivity studies established the lowest concentration of pathogen reliably detected by our assay. Ten bacterial and six fungal species were tested as 6 replicates per concentration, per analyte evaluated (**Figure 3A**). A total of 709 contrived specimens were processed resulting in an overall LoD of <25,000 CFU/mL for bacteria and <5,000 CFU/mL for fungi. Specifically, the LoDs established were 1,000 CFU/mL for *C. glabrata, C. krusei, C. parapsilosis, C. tropicalis*; 5,000 CFU/mL for *C. albicans* and *C. auris*; 7,500 CFU/mL for *E. coli, G. vaginalis, K. pneumoniae* and *P. mirabilis*; 10,000 CFU/mL for *E. faecalis; 12,500 CFU/mL for Prevotella spp.;* 15,000 CFU/mL for *A. baumanii and S. aureus;* and *25,000* CFU/mL for *B. fragilis and P. aeruginosa* (**Figure 3B**).

**Figure 3.**
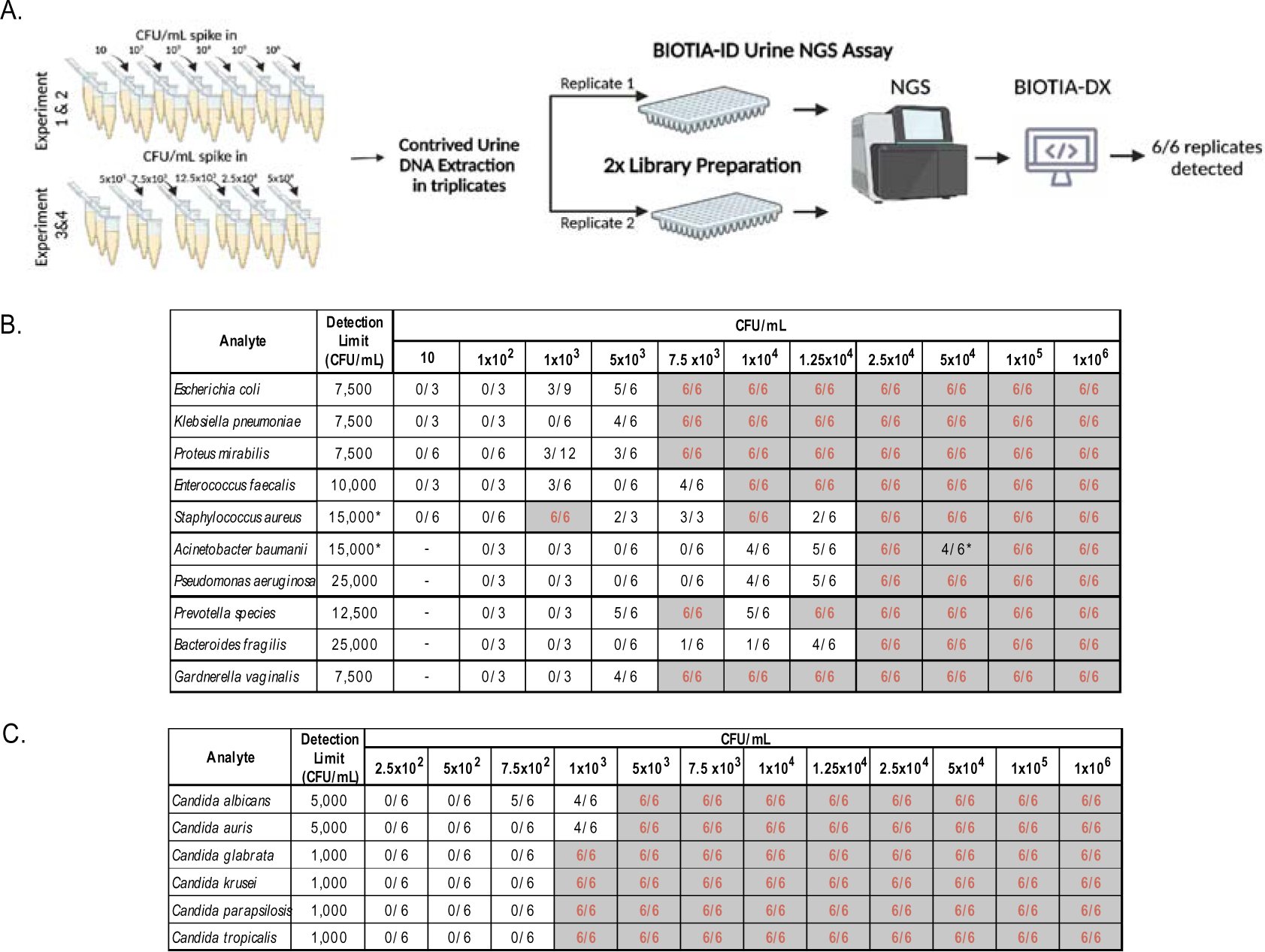
The analytical sensitivity was assessed in the most frequently found urogenital pathogens (10 bacteria and 6 fungal species) by spiking reference whole organisms into negative urine matrix. **(A)** Each pathogen was spiked in triplicate per concentration (CFU/mL) followed by genomic DNA extraction. Experiments 1 and 2 included 6 replicates for each concentration of the 10-fold dilution spike ins, ranging from 10 to 1 million CFU/mL. Experiments 3 & 4 were used to fine tune the LoD by testing 6 replicates per concentration, ranging from 5x10^3^ to 5x10^4^ CFU/mL. **(B)** Summary of replicates detected per concentration tested for each urogenital pathogen. The LoD was reproducibly verified and determined based on a 100% positivity rate resulting in an overall LoD of <25,000 CFU/mL for bacteria and <5,000 CFU/mL for fungi. Specifically, the LoDs determined for each species were 1,000 CFU/mL for *C. glabrata, C. krusei, C. parapsilosis* and *C. tropicalis*; 5,000 CFU/mL for *C. albicans* and *C. auris*; 7,500 CFU/mL for *E. coli, G. vaginalis, K. pneumoniae* and *P. mirabilis;* 10,000 CFU/mL for *E. faecalis*.; 12,500 CFU/mL for *Prevotella spp*.;15,000 CFU/mL for *A. baumanii* and *S. aureus*; 25,000 CFU/mL for *P. aeruginosa* and *B.fragilis*.

## REPRODUCIBILITY

Qualitative precision was established with inter- and intra-assay reproducibility experiments in which 19 different contrived pools containing at least 3-6 different microbial species were assessed. To account for genetic variation, a total of 66 strains belonging to 15 species of bacteria and fungi were processed. Each pool was tested in triplicate for three consecutive days obtaining a sensitivity, specificity, and qualitative reproducibility of 100%. Detailed results are presented in **Supplemental Table 2**.

## IN SILICO

The *in silico* studies assessed BIOTIA-DX’s ability to identify a large, diverse set of microorganisms absent in clinical or contrived samples and determined whether the pipeline would misidentify closely related organisms (i.e., “cross-reactivity”). The sensitivity and specificity obtained is 99.99% (a detailed breakdown of results is summarized in **Supplemental Table 3)**. Only 1.7% of synthetic samples tested (n=147) showed cross-reactivity and most species observed were within the same genus, do not have clinical relevance, or are unlikely to be found in a urine specimen (**Supplemental Table 4**). Cross-reactivities within the same genus were further differentiated by targeting specific genes containing abundant discriminating sites between species. Cross-reactive samples from 49 species were assigned the correct species in 94% of cases, further reducing false positives to 0.3% (n=27) (**Supplemental Table 4).** BIOTIA-DX overall performance on the training set, specificity, and *in silico* studies is shown on **Supplemental Figure 2**.

## ACCURACY

The accuracy verification study evaluated the precision of the assay in detecting uropathogens in clinical urine specimens compared to the current gold standard diagnostic (culture). Deidentified clinically tested urine specimens were collected from a reference laboratory based on the pathogens diagnosed by culture. A total of 335 samples (167 clinical and 168 contrived) were tested including 35 culture-negative specimens. Culture conducted by the reference laboratories reported the following CFU/mL distributions in the specimens processed: 42.5% with >100,000 CFU/mL, 11.5% with 50-100k CFU/mL, 22.4% with 10-50k CFU/mL, and 23.6% with insignificant growth or no growth.

BIOTIA-ID detected the spiked microorganisms in contrived specimens and predicted a total of 250 urogenital pathogens in the clinical specimens, classifying 83.2% (n=139) as positive for UTI (**Figure 4A, B**). Single organism infections were observed in 43.1% of the specimens while 40.1% were coinfections and polymicrobial infections (3+ pathogens, 19.8%). Conversely, 72.5% of specimens were considered single organism infections by culture (data not shown). As depicted in **Figure 4C**, BIOTIA-ID detected 27 different urogenital pathogen species, and as expected due to collection bias, higher frequencies of *E. coli, E. faecalis, K. pneumoniae, P. mirabilis* and *S. aureus* were observed. Of these, 22 additional species not reported by culture were predicted with BIOTIA-ID, representing 52.4% (n=131) of the total urogenital pathogens detected by NGS (n=250). Approximately one third (n=39) of these belonged to the 5 key bacterial species used throughout this validation, a further 29.8% (n=39) were uropathogens commonly reportable by culture (e.g., *S. agalactiae*, *Staphylococcus spp*. and other gram-negative Enterobacterales species), while 24.4% (n=32) were organisms not usually culturable with the SOC (e.g., *Aerococcus spp.*, *G. vaginalis* and *Prevotella* spp.). NGS detected *Candida* species *(C. albicans, C. glabrata, C. tropicalis and C. krusei)* in 16% (n=21) of the unreported taxa, while culture reported yeast in only 2 specimens.

**Figure 4.**
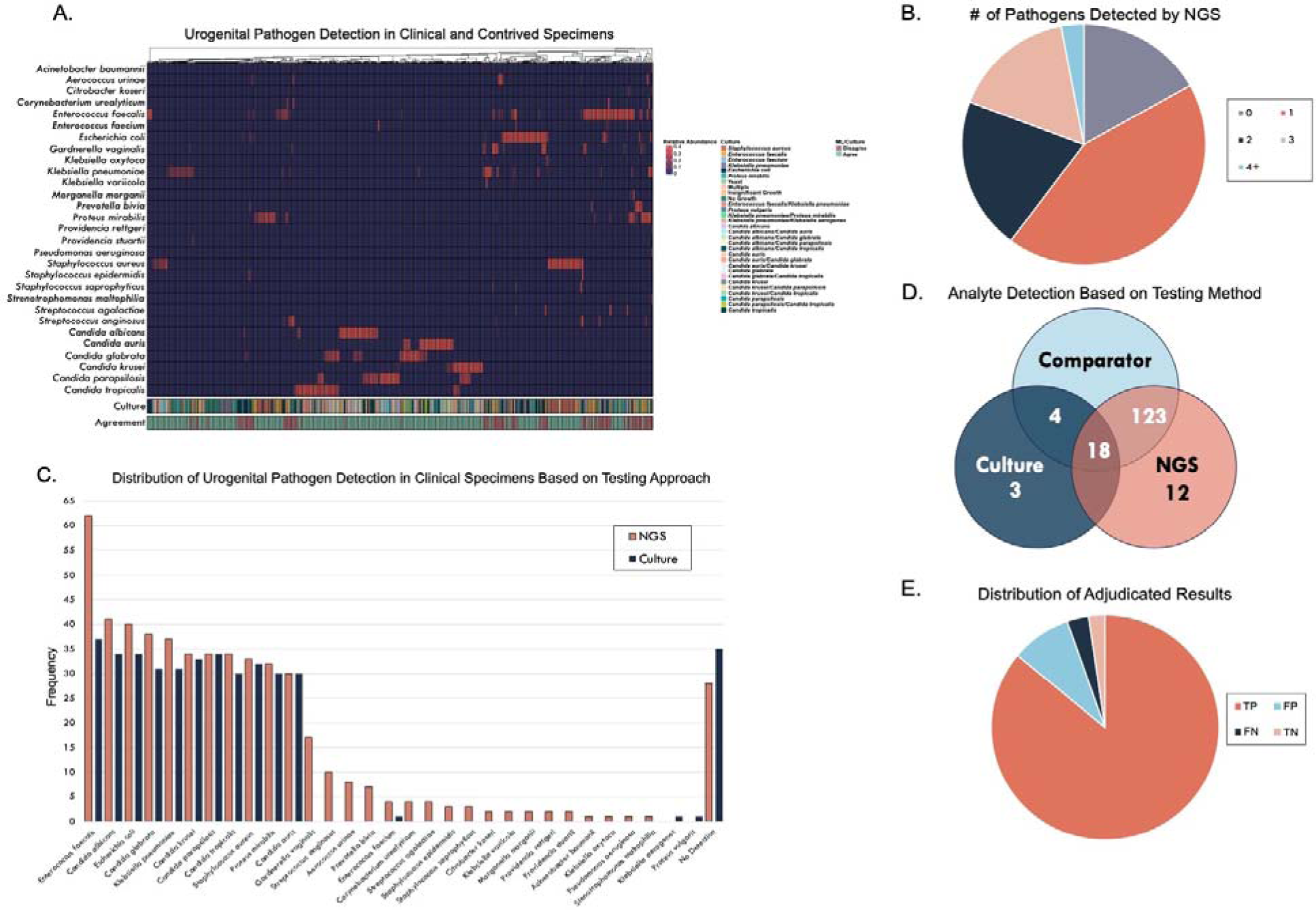
The BIOTIA-ID Urine NGS Assay accurately detects and classifies urogenital pathogens in clinical (n=167) and contrived (n=168) specimens in the accuracy study. (A) Heatmap depicts the relative abundance of taxa detected by NGS and is annotated with culture diagnosis, and agreement (green) or disagreement (purple) between BIOTIA-DX ML and culture. (B) Pie chart showing the distribution of the number of urogenital pathogens detected in clinical urine specimens tested with BIOTIA-ID. (C) Pie chart depicts the distribution of microbial analytes detected with culture (navy blue) and with NGS (pink). (D) Venn diagram illustrates the distribution and overlap of the analytes tested and detected with comparator testing for adjudicating the discrepancies found between NGS and SOC. Apparent false positive and false negative results from 19 analytes, where BIOTIA-ID differed from the original culture result (50.2% of clinical samples), were tested by qPCR or Sanger sequencing (n=123), and (E) 87% of these NGS results were found to be correct (n=107) as shown on pie chart showing the distribution of adjudicated results. TP: true positive (n=104, dark pink), TN: true negative (n=3, light pink), FP: false positive (n=12, dark blue), FN: false negative (n=4, dark gray).

Of the clinical specimens tested, 50.9% (n=85) revealed a disagreement between NGS and the gold standard and 11.3% (n=19) completely disagreed on the species detected. Due to culture limitations and the high sensitivity of NGS, comparator testing of 123 analytes (qPCR n=82, Sanger sequencing n=39) was conducted to reconcile discrepancies between findings from both methods. A subset of 19 concordant analytes were used as clinical positive controls and 100% of the analytes detected agreed across the three methods (**Figure 4D**, **Supplemental Table 5**). Apparent false positives and false negatives from 19 analytes, where BIOTIA-ID differed from the SOC, were tested and 87% of these NGS results were ultimately correct according to the comparator (**Figure 4C, D).** *E. faecalis, K. pneumoniae*, *E. coli, Candida spp., Aerococcus spp., and G. vaginalis* represented the highest frequency of disagreements (65%) between the SOC and NGS. Comparator reconciled results by taxa tested are shown in **Supplemental Figure 1**. Moreover, analytes adjudicated as false positives by NGS (n=12, 9.7%) mostly belong to *G. vaginalis* and Anginosus group streptococci, two common opportunistic pathogens of the urogenital tract. However, false negative (n=4, 3.3%) samples with analytes *E. faecalis, K. pneumoniae* and *E coli* yielded below threshold BIOTIA-ID prediction probabilities, demonstrating BIOTIA-DX’s low statistical confidence in defining the presence of the pathogen reported by the SOC.

Following comparator adjudication, the accuracy study revealed a 98.4% sensitivity and 99.7% specificity while performance in clinical specimens achieved a 97.2% sensitivity and 99.6% specificity (**Table 2**). Overall, BIOTIA-ID accomplished >99.9% sensitivity and specificity across all samples and analytes processed, exceeding ∼14.5k analytes evaluated (**Table 2**). Contrived specimens yielded >99.9% sensitivity and specificity and analyte breakdown are presented in **Supplemental Table 6.**

**Table 2.** BIOTIA-ID clinical and analytical validation performance characteristics summary by analyte category (Bacteriology, Mycology, Overall).

| Category | Method | TP | TN | FN | FP | Total | Sensitivity | Specificity |
| --- | --- | --- | --- | --- | --- | --- | --- | --- |
| Bacteriology | Clinical | 210 | 8323 | 7 | 35 | 8575 | 96.77% | 99.58% |
|  | Contrived | 1099 | 43122 | 0 | 14 | 44048 | 100.00% | 99.97% |
|  | *in silico* | 8119 | 272775 | 3 | 147 | 281044 | 99.96% | 99.95% |
|  | Total | 9431 | 332483 | 10 | 196 | 341933 | 99.89% | 99.94% |
| Mycology | Clinical | 30 | 530 | 0 | 0 | 560 | 100.00% | 100.00% |
|  | Contrived | 925 | 9240 | 1 | 2 | 10168 | 99.89% | 99.98% |
|  | *in silico* | 4142 | 41420 | 0 | 0 | 45562 | 100.00% | 100.00% |
|  | Total | 5097 | 51190 | 1 | 2 | 56290 | 99.98% | 100.00% |
| Overall | Clinical | 240 | 8853 | 7 | 35 | 9135 | 97.17% | 99.61% |
|  | Contrived | 2024 | 52362 | 1 | 16 | 54216 | 99.95% | 99.97% |
|  | *in silico* | 12261 | 314195 | 3 | 147 | 326606 | 99.98% | 99.95% |
|  | Total | 14528 | 383673 | 11 | 198 | 398223 | 99.92% | 99.95% |

## DISCUSSION

In this study, we developed and validated a clinical-grade NGS assay, BIOTIA-ID, for accurate pathogen identification in urine specimens using ML-classification. BIOTIA-ID overcomes challenges specific to urine molecular diagnostics including sample collection and transport, low gDNA yields, presence of inhibitors, high human background, and taxonomic bias due to suboptimal extraction (47–56). The laboratory process is compatible with UTT, the preservative of choice for standard urine cultures in a clinical setting. DNA extraction was optimized for efficient lysis of gram-positive bacteria and fungi ensuring sufficient gDNA yields from 2 mL of urine, specifically from low biomass samples, such as those collected from males, from catheterized patients, or from patients with prior antibiotic use. Three external controls and one internal control were implemented to validate the process (PC), monitor cross contamination coming from the process or reagents (NEC, NTC), and ensure integrity and validity of each sample minimizing false negatives due to inhibition (IPC). BIOTIA-DX has a clinically curated pathogen database, and the ML-classifier was trained with a combination of urine clinical specimens, clinical and reference isolates, and synthetic metagenomic specimens. Comparator validation of discrepant results along with a comprehensive training set increased the specificity and robustness of pathogen prediction. Analytical performance was assessed on 20 prevalent urogenital pathogens in 1,305 contrived samples, 12,250+ *in silico* simulations, and 167 clinical specimens. BIOTIA-ID demonstrated high sensitivity (99.92%) and specificity (99.95%) in predicting uropathogens, making it a promising alternative testing to culture-based diagnostics.

Although urine culture is highly reliable with reported ∼90% sensitivity and 86% specificity in healthy outpatient women, its specificity is exceptionally variable in chronically ill patients (76- 95%) and in patients with indwelling catheters where it drops close to 0%(57). BIOTIA-ID’s ability to precisely detect pathogens at low concentrations (LoD <1,000-25,000 CFU/mL) enhances its clinical utility, particularly in the context of managing high-risk immunocompromised patients, rUTI and cUTIs, and/or catheter-associated UTIs (CAUTI). UTI diagnostics currently rely on culture, and while effective, 20-30% of cases go undiagnosed (58–61), with this number increasing in complicated cases. Culture-based diagnostics fail due to numerous issues, including prior antibiotic exposure, collection or storage errors, cross-contamination, selectivity of culture conditions, and inadequate identification of fastidious, anaerobic, and fungal infections (19, 22), many of which can be addressed with NGS-based diagnostics.

In this study, half of the NGS tested clinical specimens revealed a disagreement with culture findings, with most cases detecting more than one urogenital pathogen, including the species reported by culture. About 11.3% of specimens had different species detected with NGS and about 15% exhibited clinically relevant discrepancies like detecting a gram-positive (*E. faecalis*) instead of a gram-negative pathogen (*E. coli*). Despite being highly abundant by both NGS and qPCR testing*, E. faecalis* was the top missed organism by culture. Notably, close to 43% of the clinical specimens which culture diagnosed as *P. mirabilis* (n=9/21) showed *E. faecalis* as the dominant pathogen with an NGS-based approach. *E. faecalis* is frequently under-reported by standard culture with 50% detection rates when compared to Enhanced Quantitative Urine Culture (20, 62). Further, *Candida* species were missed (n=17) or only reported by culture as yeast (n=2), representing 11.9% of the clinical specimens tested. While *Candida* can be isolated with the SOC, sensitivity is rather poor. A study compared *Candida* detection in urine samples with confirmed yeast presence on urinalysis, finding that standard urine culture identified 37% of *Candida* infections whereas using a fungal-specific media (Sabouraud Dextrose Agar) increased detection to 98%(63). Studies have shown that standard urine culture does not provide optimal growth conditions for all urogenital pathogens causing organisms such as Enterobacterales to outcompete other microbes like *Enterococcus* and *Candida* (20, 26). Failure to detect the correct pathogen results in physicians prescribing inadequate antibiotics, thus exacerbating the emergence of antimicrobial resistance, and increasing patient suffering and medical burden(64–68). For example, *Enterococcus* is the second leading cause of CAUTI, accounting for 45% of CAUTI infections and the third leading cause of hospital acquired UTIs (69–72). Although UTIs represent the primary cause for antibiotic prescriptions in people living in nursing homes, 40 to 75% of cases receive inappropriate antimicrobial therapy (1, 73–75). NGS-based metagenomic tools offer an opportunity for precise infectious disease diagnostics and have the potential to enhance antimicrobial stewardship efforts.

We preferentially procured specimens diagnosed with specific urogenital pathogens that are easily and more consistently cultured with SOC, such as *E. coli* which accounts for up to 75% of uncomplicated UTIs and 65% of cUTI cases (76–80). Standard urine culture is selective for typical microorganisms that are most commonly responsible for acute and uncomplicated UTIs; nonetheless, augmented culture methods and diagnostic tests are better suited when there is a high clinical suspicion for UTI caused by atypical organisms, as seen in neutropenia (e.g., candiduria), genitourinary tuberculosis, and urinary tract abnormalities (e.g., anaerobic bacteria) (81). Like other molecular-based studies, BIOTIA-ID showed increased detection of atypical pathogens including *Candida, G. vaginalis*, *Aerococcus spp*., anaerobic bacteria (*Prevotella spp.*) and fastidious bacteria, organisms not reported with culture-based diagnostics (30, 82–85). Although the rise of molecular based diagnostics and clinical studies using these technologies have revealed that gram-positive, atypical, or fastidious pathogens account for a larger percentage of cUTI infections, more interventional, and clinical utilization studies are needed to understand the pathogenesis and epidemiology of these understudied microbes (19, 62, 86–88). Despite specimen collection bias, BIOTIA-ID identified a significant number of atypical pathogens in the tested clinical specimens leading us to hypothesize that the detection frequency would likely rise in clinical studies that focus on culture-negative cUTI cases, a population which could greatly benefit from improved diagnostics.

Polymicrobial infections with more than two pathogens are usually considered contamination under the current SOC and reporting of these results as infection is a topic of debate (18). In accordance with other UTI studies employing molecular approaches, about 40 % of clinical specimens tested in this validation showed more than one pathogen detected (83, 85, 89–92). Most clinical algorithms designed for uropathogen detection provide optimal growth conditions for a limited number of microbes and are based on a threshold of 10^5^ CFU/mL. Understandably, many guidelines aim to prevent over prescription of antibiotics. Nonetheless, the development of improved diagnostics can help better guide antimicrobial stewardship efforts while still providing a comprehensive report. It has been demonstrated that presence of certain pathogens, even at low levels, can heighten the virulence of other dominant pathogens and could impact response to antimicrobial therapy (93–99).

Regarding concerns about the potential oversensitivity of an NGS-based assay, BIOTIA-DX was intentionally designed and trained to increase stringency and reduce false positive detection of urogenital commensals or opportunistic pathogens present at colonization levels often a common weakness of many molecular diagnostic tools and clinical studies (34, 81, 88, 100, 101). Relative abundance of microbial species, percentage of human reads, IPC detection and complexity of the microbial profiles are features implemented to enhance the BIOTIA-DX’s ability to correctly predict an infection. This study highlights how ML coupled with extensive laboratory and bioinformatic validation could improve infectious disease diagnostics by addressing the failures of culture-based diagnostics. It is worth noting that we observed a strong linear correlation between spiked CFU/mL and relative abundance of LoD contrived samples (data not shown). The complexity of a clinical urine specimen requires the consideration and development of additional pipeline features for the implementation of a quantification metric that could be relevant and useful to physicians. Additional clinical studies are required to generate a robust dataset, facilitating the creation of an updated, user-friendly NGS-based metric that aligns more closely with current clinical and antibiotic stewardship guidelines for UTI treatment and management (17, 28, 85, 102) Further studies and real-world evaluations are warranted to validate the clinical impact and cost-effectiveness of BIOTIA-ID as a UTI diagnostic tool. The observed performance characteristics suggest potential clinical applications where the advantages of sensitivity, specificity, and comprehensive testing outweigh some of the limitations associated with sequencing cost and TAT. Some use cases that would most benefit from this approach include symptomatic patients with negative culture, cUTI and rUTI, and immunocompromised patients with nonspecific infection symptoms who are at high-risk of developing sepsis.

BIOTIA-ID represents a significant advancement in UTI diagnostics, offering notable advantages over culture-based methods. Its high accuracy, low LoD, and potential to guide targeted antimicrobial therapy support the goals of antimicrobial stewardship. Implementation of BIOTIA-ID in clinical settings has the potential to improve patient outcomes, reduce the misuse of antibiotics, and contribute to the global fight against antimicrobial resistance.

## Data Availability

All data produced in the present study are available upon reasonable request to the authors.

## ACKNOWLEDGEMENTS

We thank Zaineb Bello for clinical specimen collection and laboratory operation support; Tiara Rivera and Tamara Goncharuk for sequencing support; Yehudah Gruenstein and LabQ Team for clinical specimen collection; Adam Nagyhazy-Horvath, Eszter Szollosy, Patrik Blik, the Abesse Team, Pierre Davidoff, Cory Mason for IT support; Taylor Paisie for bioinformatic support; Talisa Vega-Kline for logistical support.

The BIOTIA-DX software pipeline work used Cromwell On Azure; the Microsoft Genomics supported implementation of the Broad Institute’s Cromwell workflow engine on Azure. The development and validation work used the Extreme Science and Engineering Discovery Environment (XSEDE), which is supported by National Science Foundation grant (ACI- 1548562). Specifically, it used the Bridges system, which is supported by NSF award (ACI- 1445606), at the Pittsburgh Supercomputing Center (PSC).

---

**Supplemental Figure 1.**
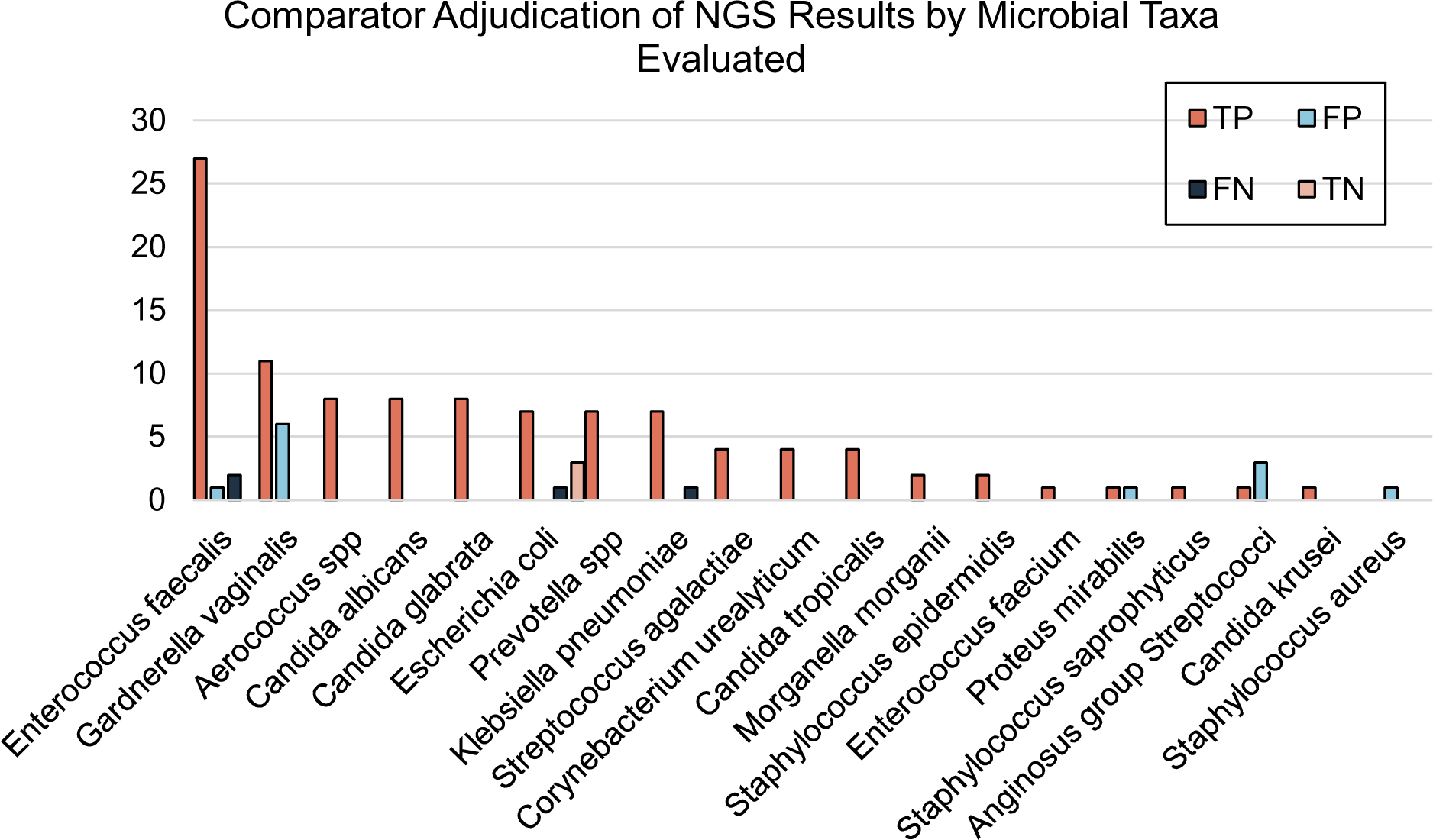
Genomic DNA from urine specimens with apparent false positive and false negative results where BIOTIA-ID differed from the original culture result (49.2% of analytes detected), were tested by qPCR or Sanger sequencing (n=123). 87% of NGS results were ultimately found to be correct by the orthogonal adjudication. Bar chart shows comparator adjudicated results for the urogenital pathogens tested with comparator assays. TP: true positive (dark pink), FP: false positive (dark blue), FN: false negative (gray), TN: true negative (light pink).

**Supplemental Figure 2.**
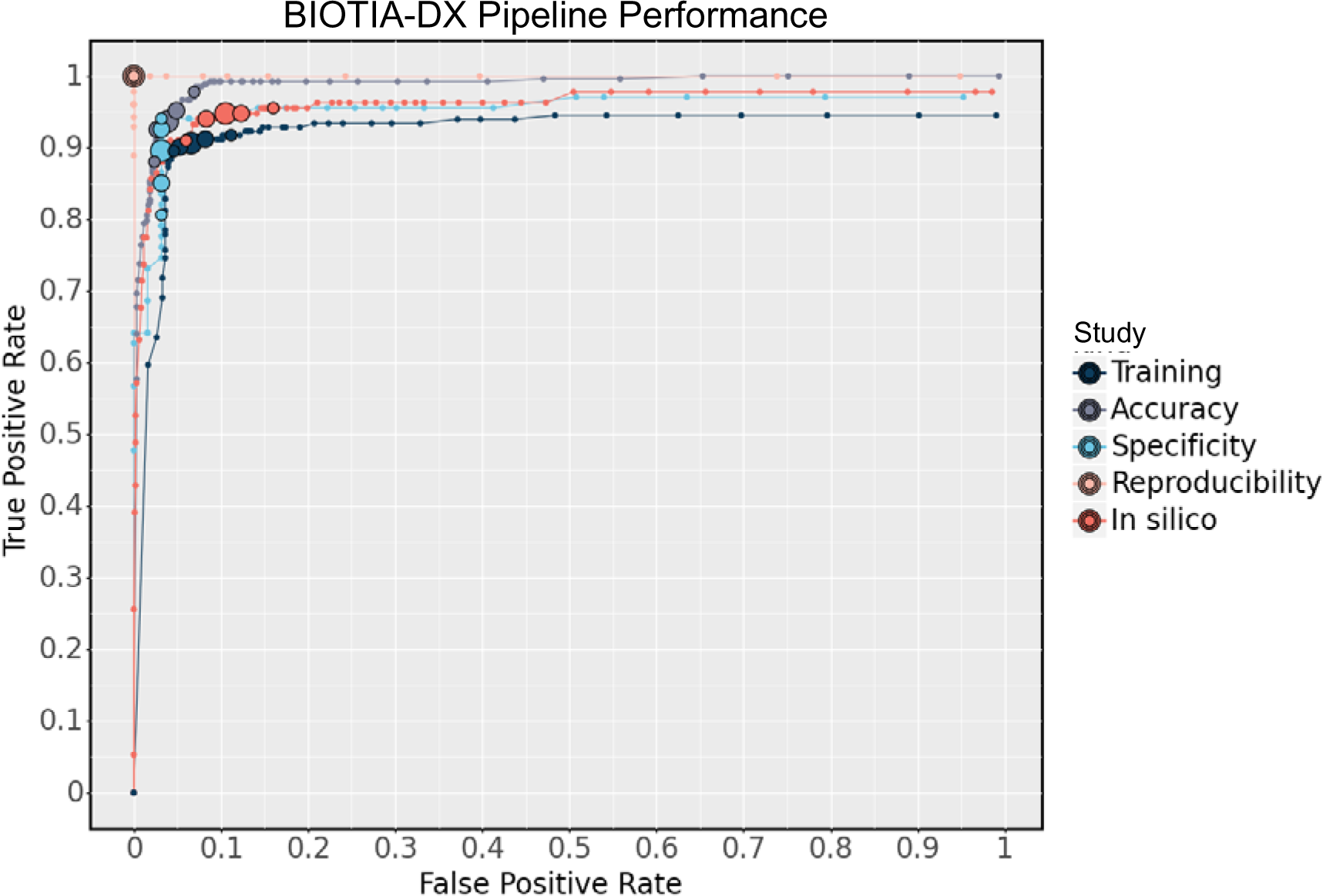
BIOTIA-DX ML Performance in samples processed on the training (dark blue), *in silico* (dark pink), accuracy (gray), reproducibility (light pink) and specificity (light blue) studies.

