## Supplemental Tables for "Analytical Validation of a Highly Accurate and Reliable Next-Generation Sequencing-Based Urine Assay"

**Supplemental Table 1.** Summary of comparator assays used in this study

| Taxa | Target gene | Assay Type | Forward Primer Sequence | Reverse Primer Sequence | Probe Sequence | Reference |
|---|---|---|---|---|---|---|
| *Escherichia coli* | uidA | Taqman | CGGAAGCAACGCGTAAACTC | TGAGCGTCGCAGAACATTACA | CGCGTCCGATCACCTGCGTC | Silkie, et al., 2008 |
| *Klebsiella pneumoniae* | khe | Taqman | GATGAAACGACCTGATTGCATTC | CCGGGCTGTCGGGATAAG | GCGAACTGGAAGGGCCCG | Hartman, et al., 2009 |
| *Proteus mirabilis* | ureR | Taqman | CCATCAGATTATGTCATTCAA | GAGGAAAATGCAATTTATCTTTA | CACACCCTACCCAACATTCATTTCA | Liu, et al., 2019 |
| *Enterococcus faecalis* | 16S | Taqman | CGCTTCTTTCCTCCCGAGT | GCCATGCGGCATAAACTG | CAATTGGAAAGAGGAGTGGCGGACG | Santo Domingo et al., 2003 |
| *Staphylococcus aureus* | ebpS | Taqman | CCACATGCCTCTAATAATG | GCGATTTTATTTTCTTTTGTAC | ATGCCATGCCTCCAAATATCGC | Liu, et al., 2019 |
| *Gardnerella vaginalis* | cpn60 | Taqman | CGCATCTGCTAAGGATGTTG | CAGCAATCTTTTCGCCAACT | TGCAACTATTTCTGCAGCAGATCC | Menard et al., 2008 |
| *Aerococcus urinae* | 16S | Taqman | GACGGCTTTGCTGTCACTTATCG | GCTATGCATCATTGSCTTGGTAG | TGCATTAGCTCGTTGGTGGG | Biotia |
| *Prevotella species* | 16S | SYBR green | GGTTCTGAGAGGAAGGTCCCC | TCCTGCACGCTACTTGGCTG | NA | Stevenson and Weimer, 2007 |
| *Candida species* | ITS | Sanger | TCCGTAGGTGAACCTGCGG | TCCTCCGCTTATTGATATGC | NA | White et al., 1990 |
| *Anaerococcus vaginalis* | 16S | Sanger | TGATTTCTTCGGAATGAAATTAAGTGGATTA | GGTCATTTTATCATGCGATATTTTGACTTT | NA | Biotia |
| *Corynebacterium urealyticum* | 16S | Sanger | CCTGCTTGCAGGGTACTCGA | CACCAACCACACTAAAGATTGGTC | NA | Biotia |
| *Enterococcus faecium* | 16S | Sanger | TTCTTTTTCCACCGGAGCTT | AACCATGCGGTTTYGATTG | NA | Ryu et al., 2013 |
| *Morganella morganii* | 16S | Sanger | AGCTTGCTTCTCTGCTGACGAG | GAGGCCCGAAGGTCCCCCG | NA | Biotia |
| *Staphylococcus epidermidis* | 16S | Sanger | GTTCAATAGTGAAAGACGGTTTTGCTGTC | GTTTTACGATCCGAAGACCTTCATCAC | NA | Biotia |
| *Staphylococcus saprophyticus* | 16S | Sanger | TAAAGTGAAAGATGGTTTTGCTATC | GTTTTACGAGCCGAAACCCTTCATCAC | NA | Biotia |
| *Streptococcus agalactiae* | 16S | Sanger | KGTTTGGTGTTTACACTAGACTG | TTACCGTCACTTGGTAGATTTTCCA | NA | Biotia |
| *Streptococcus anginosus* | 16S | Sanger | CGTAGTTTACTACACCGTATTCTGTGA | GTACCGTCACTGTGTGAAC | NA | Biotia |

**Supplementary Table 2A.** Summary of Replicates Detected in the Reproducibility Studies

| Microbial Species | Replicates Detected | Qualitative Reproducibility |
|---|---|---|
| *Enterococcus faecalis* | 81/81 | 100% |
| *Escherichia coli* | 81/81 | 100% |
| *Klebsiella pneumoniae* | 81/81 | 100% |
| *Proteus mirabilis* | 81/81 | 100% |
| *Staphylococcus aureus* | 81/81 | 100% |
| *Candida albicans* | 45/45 | 100% |
| *Candida auris* | 45/45 | 100% |
| *Candida glabrata* | 45/45 | 100% |
| *Candida krusei* | 45/45 | 100% |
| *Candida parapsilosis* | 45/45 | 100% |
| *Candida tropicalis* | 45/45 | 100% |
| *Candida dubliniensis* | 9/9 | 100% |
| *Candida guillermondii* | 9/9 | 100% |
| *Candida kefyr* | 9/9 | 100% |
| *Candida lusitaniae* | 9/9 | 100% |
| *Negative* | 42/42 | 100% |

**Supplementary Table 2B.** Reproducibility Studies Results by Concentration

| Analyte | Strain | Load | Day 1 | Day 2 | Day 3 | Total |
|---|---|---|---|---|---|---|
| *Enterococcus faecalis* | URV-032-2 | 15,000 | 3/3 | 3/3 | 3/3 | 9/9 |
| *Escherichia coli* | URV-012 | 15,000 | 3/3 | 3/3 | 3/3 | 9/9 |
| *Klebsiella pneumoniae* | URV-155-1 | 15,000 | 3/3 | 3/3 | 3/3 | 9/9 |
| *Proteus mirabilis* | URV-023 | 15,000 | 3/3 | 3/3 | 3/3 | 9/9 |
| *Staphylococcus aureus* | ATCC-43300 | 15,000 | 3/3 | 3/3 | 3/3 | 9/9 |
| *Enterococcus faecalis* | URV-032-2 | 20,000 | 3/3 | 3/3 | 3/3 | 9/9 |
| *Escherichia coli* | URV-012 | 20,000 | 3/3 | 3/3 | 3/3 | 9/9 |
| *Klebsiella pneumoniae* | URV-155-1 | 20,000 | 3/3 | 3/3 | 3/3 | 9/9 |
| *Proteus mirabilis* | URV-023 | 20,000 | 3/3 | 3/3 | 3/3 | 9/9 |
| *Staphylococcus aureus* | ATCC-43300 | 20,000 | 3/3 | 3/3 | 3/3 | 9/9 |
| *Enterococcus faecalis* | URV-032-2 | 25,000 | 3/3 | 3/3 | 3/3 | 9/9 |
| *Escherichia coli* | URV-012 | 25,000 | 3/3 | 3/3 | 3/3 | 9/9 |
| *Klebsiella pneumoniae* | URV-155-1 | 25,000 | 3/3 | 3/3 | 3/3 | 9/9 |
| *Proteus mirabilis* | URV-023 | 25,000 | 3/3 | 3/3 | 3/3 | 9/9 |
| *Staphylococcus aureus* | ATCC-43300 | 25,000 | 3/3 | 3/3 | 3/3 | 9/9 |
| *Enterococcus faecalis* | URV-220 | 25,000 | 3/3 | 3/3 | 3/3 | 9/9 |
| *Escherichia coli* | URV-005-1 | 25,000 | 3/3 | 3/3 | 3/3 | 9/9 |
| *Klebsiella pneumoniae* | URV-038 | 25,000 | 3/3 | 3/3 | 3/3 | 9/9 |
| *Proteus mirabilis* | URV-010 | 25,000 | 3/3 | 3/3 | 3/3 | 9/9 |
| *Staphylococcus aureus* | SJF-038 | 25,000 | 3/3 | 3/3 | 3/3 | 9/9 |
| *Enterococcus faecalis* | URV-032-2 | 25,000 | 3/3 | 3/3 | 3/3 | 9/9 |
| *Escherichia coli* | URV-012 | 25,000 | 3/3 | 3/3 | 3/3 | 9/9 |
| *Klebsiella pneumoniae* | URV-036 | 25,000 | 3/3 | 3/3 | 3/3 | 9/9 |
| *Proteus mirabilis* | URV-014 | 25,000 | 3/3 | 3/3 | 3/3 | 9/9 |
| *Staphylococcus aureus* | SJF-044 | 25,000 | 3/3 | 3/3 | 3/3 | 9/9 |
| *Enterococcus faecalis* | URV-007 | 25,000 | 3/3 | 3/3 | 3/3 | 9/9 |
| *Escherichia coli* | URV-029 | 25,000 | 3/3 | 3/3 | 3/3 | 9/9 |
| *Klebsiella pneumoniae* | URV-022 | 25,000 | 3/3 | 3/3 | 3/3 | 9/9 |
| *Proteus mirabilis* | URV-040 | 25,000 | 3/3 | 3/3 | 3/3 | 9/9 |
| *Staphylococcus aureus* | SJF-034 | 25,000 | 3/3 | 3/3 | 3/3 | 9/9 |
| *Enterococcus faecalis* | URV-026 | 25,000 | 3/3 | 3/3 | 3/3 | 9/9 |
| *Escherichia coli* | URV-006 | 25,000 | 3/3 | 3/3 | 3/3 | 9/9 |
| *Klebsiella pneumoniae* | URV-031 | 25,000 | 3/3 | 3/3 | 3/3 | 9/9 |
| *Proteus mirabilis* | URV-060 | 25,000 | 3/3 | 3/3 | 3/3 | 9/9 |
| *Staphylococcus aureus* | SJF-035 | 25,000 | 3/3 | 3/3 | 3/3 | 9/9 |
| *Enterococcus faecalis* | URV-261 | 25,000 | 3/3 | 3/3 | 3/3 | 9/9 |
| *Escherichia coli* | URV-021 | 25,000 | 3/3 | 3/3 | 3/3 | 9/9 |
| *Klebsiella pneumoniae* | URV-155-1 | 25,000 | 3/3 | 3/3 | 3/3 | 9/9 |
| *Proteus mirabilis* | URV-016 | 25,000 | 3/3 | 3/3 | 3/3 | 9/9 |
| *Staphylococcus aureus* | SJF-041 | 25,000 | 3/3 | 3/3 | 3/3 | 9/9 |
| *Enterococcus faecalis* | URV-032-2 | 100,000 | 3/3 | 3/3 | 3/3 | 9/9 |
| *Escherichia coli* | AR-104 | 100,000 | 3/3 | 3/3 | 3/3 | 9/9 |
| *Klebsiella pneumoniae* | AR-557 | 100,000 | 3/3 | 3/3 | 3/3 | 9/9 |
| *Proteus mirabilis* | AR-156 | 100,000 | 3/3 | 3/3 | 3/3 | 9/9 |
| *Staphylococcus aureus* | SJF-035 | 100,000 | 3/3 | 3/3 | 3/3 | 9/9 |
| *Candida albicans* | 60193 | 25,000 | 3/3 | 3/3 | 3/3 | 9/9 |
| *Candida auris* | MYA-5000 | 25,000 | 3/3 | 3/3 | 3/3 | 9/9 |
| *Candida glabrata* | 15126 | 25,000 | 3/3 | 3/3 | 3/3 | 9/9 |
| *Candida krusei* | 14243 | 25,000 | 3/3 | 3/3 | 3/3 | 9/9 |
| *Candida parapsilosis* | 22019 | 25,000 | 3/3 | 3/3 | 3/3 | 9/9 |
| *Candida tropicalis* | 1369 | 25,000 | 3/3 | 3/3 | 3/3 | 9/9 |

| | | | | | | |
|---|---|---|---|---|---|---|
| *Candida albicans* | 10231 | 25,000 | 3/3 | 3/3 | 3/3 | 9/9 |
| *Candida auris* | MYA-5001 | 25,000 | 3/3 | 3/3 | 3/3 | 9/9 |
| *Candida glabrata* | 2001 | 25,000 | 3/3 | 3/3 | 3/3 | 9/9 |
| *Candida krusei* | 34135 | 25,000 | 3/3 | 3/3 | 3/3 | 9/9 |
| *Candida parapsilosis* | 90018 | 25,000 | 3/3 | 3/3 | 3/3 | 9/9 |
| *Candida tropicalis* | 1369 | 25,000 | 3/3 | 3/3 | 3/3 | 9/9 |
| *Candida albicans* | 14053 | 25,000 | 3/3 | 3/3 | 3/3 | 9/9 |
| *Candida auris* | MYA-5002 | 25,000 | 3/3 | 3/3 | 3/3 | 9/9 |
| *Candida glabrata* | 66032 | 25,000 | 3/3 | 3/3 | 3/3 | 9/9 |
| *Candida krusei* | 90878 | 25,000 | 3/3 | 3/3 | 3/3 | 9/9 |
| *Candida parapsilosis* | SJF-019 | 25,000 | 3/3 | 3/3 | 3/3 | 9/9 |
| *Candida tropicalis* | 750 | 25,000 | 3/3 | 3/3 | 3/3 | 9/9 |
| *Candida albicans* | 90029 | 25,000 | 3/3 | 3/3 | 3/3 | 9/9 |
| *Candida auris* | MYA-5003 | 25,000 | 3/3 | 3/3 | 3/3 | 9/9 |
| *Candida glabrata* | 90030 | 25,000 | 3/3 | 3/3 | 3/3 | 9/9 |
| *Candida krusei* | 6258 | 25,000 | 3/3 | 3/3 | 3/3 | 9/9 |
| *Candida parapsilosis* | SJF-032 | 25,000 | 3/3 | 3/3 | 3/3 | 9/9 |
| *Candida tropicalis* | 13803 | 25,000 | 3/3 | 3/3 | 3/3 | 9/9 |
| *Candida albicans* | URV-018 | 25,000 | 3/3 | 3/3 | 3/3 | 9/9 |
| *Candida auris* | CDC B11903 | 25,000 | 3/3 | 3/3 | 3/3 | 9/9 |
| *Candida glabrata* | MYA-2950 | 25,000 | 3/3 | 3/3 | 3/3 | 9/9 |
| *Candida krusei* | URV-019 | 25,000 | 3/3 | 3/3 | 3/3 | 9/9 |
| *Candida parapsilosis* | SJF-033 | 25,000 | 3/3 | 3/3 | 3/3 | 9/9 |
| *Candida tropicalis* | 66029 | 25,000 | 3/3 | 3/3 | 3/3 | 9/9 |
| *Candida dubliniensis* | 3949 | 25,000 | 3/3 | 3/3 | 3/3 | 9/9 |
| *Candida guillermondii* | 6260 | 25,000 | 3/3 | 3/3 | 3/3 | 9/9 |
| *Candida kefyr* | 66028 | 25,000 | 3/3 | 3/3 | 3/3 | 9/9 |
| *Candida lusitaniae* | 34449 | 25,000 | 3/3 | 3/3 | 3/3 | 9/9 |
| Negative Samples | - | - | 14/14 | 14/14 | 14/14 | 42/42 |

**Supplementary Table 3A.** Performance Characteristics of in silico Studies per Key Urogenital Taxa

| Key pathogen | Method | TP | TN | FN | FP | Total | Sensitivity | Specificity |
|---|---|---|---|---|---|---|---|---|
| *Escherichia coli* | Clinical | | | | | | | |
| | Contrived | | | | | | | |
| | *in silico* | 3 | 8239 | 0 | 24 | 8266 | 100.0% | 99.7% |
| | *Total* | | | | | | | |
| *Klebsiella pneumoniae* | Clinical | | | | | | | |
| | Contrived | | | | | | | |
| | *in silico* | 3 | 8250 | 0 | 13 | 8266 | 100.0% | 99.8% |
| | *Total* | | | | | | | |
| *Proteus mirabilis* | Clinical | | | | | | | |
| | Contrived | | | | | | | |
| | in silico | 3 | 8255 | 0 | 8 | 8266 | 100.0% | 99.9% |
| | *Total* | | | | | | | |
| *Enterococcus faecalis* | Clinical | | | | | | | |
| | Contrived | | | | | | | |
| | in silico | 3 | 8263 | 0 | 0 | 8266 | 100.0% | 100.0% |
| | *Total* | | | | | | | |
| *Staphylococcus aureus* | Clinical | | | | | | | |
| | Contrived | | | | | | | |
| | *in silico* | 3 | 8252 | 0 | 11 | 8266 | 100.0% | 99.9% |
| | *Total* | | | | | | | |
| *Citrobacter species* | Clinical | | | | | | | |
| | Contrived | | | | | | | |
| | in silico | 20 | 8246 | 0 | 0 | 8266 | 100.0% | 100.0% |
| | *Total* | | | | | | | |
| *Enterobacter aerogenes* | Clinical | | | | | | | |
| | Contrived | | | | | | | |
| | in silico | 3 | 8263 | 0 | 0 | 8266 | 100.0% | 100.0% |
| | *Total* | | | | | | | |
| *Enterobacter cloacae* | Clinical | | | | | | | |
| | Contrived | | | | | | | |
| | *in silico* | 3 | 8253 | 0 | 10 | 8266 | 100.0% | 99.9% |
| | *Total* | | | | | | | |
| *Klebsiella variicola* | Clinical | | | | | | | |
| | Contrived | | | | | | | |
| | in silico | 3 | 8260 | 0 | 3 | 8266 | 100.0% | 100.0% |
| | *Total* | | | | | | | |
| *Klebsiella oxytoca* | Clinical | | | | | | | |
| | Contrived | | | | | | | |
| | *in silico* | 3 | 8260 | 0 | 3 | 8266 | 100.0% | 100.0% |
| | *Total* | | | | | | | |
| *Morganella morganii* | Clinical | | | | | | | |
| | Contrived | | | | | | | |
| | in silico | 3 | 8262 | 0 | 1 | 8266 | 100.0% | 100.0% |
| | *Total* | | | | | | | |
| *Proteus vulgaris* | Clinical | | | | | | | |
| | Contrived | | | | | | | |
| | *in silico* | 3 | 8256 | 0 | 7 | 8266 | 100.0% | 99.9% |
| | *Total* | | | | | | | |
| *Providencia rettgeri* | Clinical | | | | | | | |
| | Contrived | | | | | | | |
| | *in silico* | 3 | 8260 | 0 | 3 | 8266 | 100.0% | 100.0% |
| | *Total* | | | | | | | |
| *Providencia stuartii* | Clinical | | | | | | | |
| | Contrived | | | | | | | |
| | *in silico* | 3 | 8260 | 0 | 3 | 8266 | 100.0% | 100.0% |
| | *Total* | | | | | | | |
| *Raoultella ornithinolytica* | Clinical | | | | | | | |
| | Contrived | | | | | | | |
| | *in silico* | 3 | 8260 | 0 | 3 | 8266 | 100.0% | 100.0% |
| | *Total* | | | | | | | |
| *Serratia marcescens* | Clinical | | | | | | | |
| | Contrived | | | | | | | |
| | *in silico* | 3 | 8245 | 0 | 18 | 8266 | 100.0% | 99.8% |
| | *Total* | | | | | | | |
| *Acinetobacter baumanii* | Clinical | | | | | | | |
| | Contrived | | | | | | | |
| | in silico | 3 | 8244 | 0 | 19 | 8266 | 100.0% | 99.8% |
| | *Total* | | | | | | | |

| | | | | | | | | |
|---|---|---|---|---|---|---|---|---|
| *Acinetobacter lwolfii* | Clinical | | | | | | | |
| | Contrived | | | | | | | |
| | in silico | 3 | 8260 | 0 | 3 | 8266 | 100.0% | 100.0% |
| | *Total* | | | | | | | |
| *Pseudomonas aeruginosa* | Clinical | | | | | | | |
| | Contrived | | | | | | | |
| | *in silico* | 3 | 8260 | 0 | 3 | 8266 | 100.0% | 100.0% |
| | *Total* | | | | | | | |
| *Strenotrophomonas maltophilia* | Clinical | | | | | | | |
| | Contrived | | | | | | | |
| | *in silico* | 3 | 8252 | 0 | 11 | 8266 | 100.0% | 99.9% |
| | *Total* | | | | | | | |
| *Aerococcus species* | Clinical | | | | | | | |
| | Contrived | | | | | | | |
| | *in silico* | 21 | 8242 | 3 | 0 | 8266 | 87.5% | 100.0% |
| | *Total* | | | | | | | |
| Anginosus group Streptococci | Clinical | | | | | | | |
| | Contrived | | | | | | | |
| | *in silico* | 5 | 8261 | 0 | 0 | 8266 | 100.0% | 100.0% |
| | *Total* | | | | | | | |
| *Streptococcus agalactiae* | Clinical | | | | | | | |
| | Contrived | | | | | | | |
| | in silico | 3 | 8263 | 0 | 0 | 8266 | 100.0% | 100.0% |
| | *Total* | | | | | | | |
| *Enterococcus faecium* | Clinical | | | | | | | |
| | Contrived | | | | | | | |
| | in silico | 3 | 8260 | 0 | 3 | 8266 | 100.0% | 100.0% |
| | *Total* | | | | | | | |
| *Corynebacterium urealyticum* | Clinical | | | | | | | |
| | Contrived | | | | | | | |
| | in silico | 3 | 8263 | 0 | 0 | 8266 | 100.0% | 100.0% |
| | *Total* | | | | | | | |
| *Staphylococcus epidermidis* | Clinical | | | | | | | |
| | Contrived | | | | | | | |
| | *in silico* | 3 | 8263 | 0 | 0 | 8266 | 100.0% | 100.0% |
| | *Total* | | | | | | | |
| *Staphylococcus lugdunensis* | Clinical | | | | | | | |
| | Contrived | | | | | | | |
| | *in silico* | 3 | 8263 | 0 | 0 | 8266 | 100.0% | 100.0% |
| | *Total* | | | | | | | |
| *Staphylococcus saprophyticus* | Clinical | | | | | | | |
| | Contrived | | | | | | | |
| | *in silico* | 3 | 8263 | 0 | 0 | 8266 | 100.0% | 100.0% |
| | *Total* | | | | | | | |
| Mitis group Streptococci | Clinical | | | | | | | |
| | Contrived | | | | | | | |
| | in silico | 3 | 8262 | 0 | 1 | 8266 | 100.0% | 100.0% |
| | *Total* | | | | | | | |
| *Anaerococcus vaginalis* | Clinical | | | | | | | |
| | Contrived | | | | | | | |
| | *in silico* | 3 | 8263 | 0 | 0 | 8266 | 100.0% | 100.0% |
| | *Total* | | | | | | | |
| *Bacteroides fragilis* | Clinical | | | | | | | |
| | Contrived | | | | | | | |
| | *in silico* | 3 | 8263 | 0 | 0 | 8266 | 100.0% | 100.0% |
| | *Total* | | | | | | | |
| *Prevotella species* | Clinical | | | | | | | |
| | Contrived | | | | | | | |
| | *in silico* | 9 | 8257 | 0 | 0 | 8266 | 100.0% | 100.0% |
| | *Total* | | | | | | | |
| *Gardnerella vaginalis* | Clinical | | | | | | | |
| | Contrived | | | | | | | |
| | *in silico* | 3 | 8263 | 0 | 0 | 8266 | 100.0% | 100.0% |
| | *Total* | | | | | | | |
| Negative Organisms* | Contrived | | | | | | | |
| | *in silico* | 7977 | 289 | 0 | 0 | 8266 | 100.0% | 100.0% |
| | *Total* | | | | | | | |

Supplementary Table 3B. Performance Characteristics of in silico Studies per Key Fungal Urogenital Taxa

| Key pathogen | Method | TP | TN | FN | FP | Total | Sensitivity | Specificity |
|---|---|---|---|---|---|---|---|---|
| *Candida albicans* | Clinical | | | | | | | |
| | Contrived | | | | | | | |
| | *in silico* | 9 | 4133 | 0 | 0 | 4142 | 100.0% | 100.0% |
| | *Total* | | | | | | | |
| *Candida auris* | Clinical | | | | | | | |
| | Contrived | | | | | | | |
| | *in silico* | 9 | 4133 | 0 | 0 | 4142 | 100.0% | 100.0% |
| | *Total* | | | | | | | |
| *Candida dubliniensis* | Clinical | | | | | | | |
| | Contrived | | | | | | | |
| | *in silico* | 9 | 4133 | 0 | 0 | 4142 | 100.0% | 100.0% |
| | *Total* | | | | | | | |
| *Candida glabrata* | Clinical | | | | | | | |
| | Contrived | | | | | | | |
| | *in silico* | 9 | 4133 | 0 | 0 | 4142 | 100.0% | 100.0% |
| | *Total* | | | | | | | |
| *Candida guilliermondii* | Clinical | | | | | | | |
| | Contrived | | | | | | | |
| | *in silico* | 9 | 4133 | 0 | 0 | 4142 | 100.0% | 100.0% |
| | *Total* | | | | | | | |
| *Candida kefyr* | Clinical | | | | | | | |
| | Contrived | | | | | | | |
| | *in silico* | 9 | 4133 | 0 | 0 | 4142 | 100.0% | 100.0% |
| | *Total* | | | | | | | |
| *Candida krusei* | Clinical | | | | | | | |
| | Contrived | | | | | | | |
| | *in silico* | 9 | 4133 | 0 | 0 | 4142 | 100.0% | 100.0% |
| | *Total* | | | | | | | |
| *Candida lusitaniae* | Clinical | | | | | | | |
| | Contrived | | | | | | | |
| | *in silico* | 9 | 4133 | 0 | 0 | 4142 | 100.0% | 100.0% |
| | *Total* | | | | | | | |
| *Candida parapsilopsis* | Clinical | | | | | | | |
| | Contrived | | | | | | | |
| | *in silico* | 9 | 4133 | 0 | 0 | 4142 | 100.0% | 100.0% |
| | *Total* | | | | | | | |
| *Candida tropicalis* | Clinical | | | | | | | |
| | Contrived | | | | | | | |
| | *in silico* | 9 | 4133 | 0 | 0 | 4142 | 100.0% | 100.0% |
| | *Total* | | | | | | | |
| Negative Organisms* | Contrived | | | | | | | |
| | *in silico* | 4052 | 90 | 0 | 0 | 4142 | 100.0% | 100.0% |
| | *Total* | | | | | | | |

**Supplementary Table 4.** Summary of *in silico* study cross reactive species

| Analyte | # of samples | Cross reactive species |
|---|---|---|
| *Acinetobacter baumanii* | 19 | *Acinetobacter genomosp* |
| *Acinetobacter lwoffii* | 3 | *Acinetobacter pseudolwoffii* |
| *Enterobacter cloacae* | 10 | *Enterobacter chengduensis* |
| *Enterococcus faecium* | 3 | *Enterococcus lactis* |
| *Escherichia coli* | 48 | *Escherichia marmotae* |
| *Klebsiella oxytoca* | 3 | *Klebsiella grimontii* |
| *Klebsiella pneumoniae* | 13 | *Klebsiella quasipneumoniae* |
| *Klebsiella variicola* | 3 | *Klebsiella quasivariicola* |
| *Morganella morganii* | 1 | *Morganella psychrotolerans* |
| *Proteus mirabilis* | 8 | *Proteus penneri* |
| *Proteus vulgaris* | 7 | *Proteus faecis* |
| *Providencia rettgeri* | 3 | *Providencia huaxiensis* |
| *Providencia stuartii* | 3 | *Providencia thailandensis* |
| *Pseudomonas aeruginosa* | 3 | *Pseudomonas paraeruginosa* |
| *Raoultella ornithinolytica* | 3 | *Raoultella planticola* |
| *Serratia marcescens* | 18 | *Serratia nematodiphila* |
| *Staphylococcus aureus* | 11 | *Staphylococcus roterodami* |
| *Stenotrophomonas maltophilia* | 11 | *Stenotrophomonas indicatrix* |
| *Streptococcus mitis* | 1 | *Streptococcus pseudopneumoniae* |
| **Simulated Reads Species** | **# of samples** | **Cross reactive species** |
| *Proteus vulgaris* | 2 | *Proteus terrae* |
| *Serratia nematodiphila* | 1 | *Serratia ureilytica* |
| *Streptococcus pseudopneumoniae* | 1 | *Streptococcus pneumoniae* |
| *Staphylococcus singaporensis* | 3 | *Staphylococcus haemolyticus* |

**Supplementary Table 5.** Comparator testing was used to resolve discrepancies between the BIOTIA-DX and culture. A combination of qPCR assays and Sanger sequencing were used for resolving such discrepancies. Assays are summarized in Supplementary Table 1.

**qPCR Comparator Testing Results**

| Sample | Analyte | Prediction Probability | Relative abundance | Biotia-ID Result | Comparator result | qPCR Ct value | Outcome |
|---|---|---|---|---|---|---|---|
| URV-104 | *Enterococcus faecalis* | 0.988 | 0.198 | Detected | Detected | 16.07 | True Positive |
| URV-110 | *Enterococcus faecalis* | 0.999 | 0.135 | Detected | Detected | 15.79 | True Positive |
| URV-118 | *Enterococcus faecalis* | 0.999 | 0.215 | Detected | Detected | 24.00 | True Positive |
| URV-122 | *Enterococcus faecalis* | 0.972 | 0.081 | Detected | Detected | 16.76 | True Positive |
| URV-127 | *Enterococcus faecalis* | 0.998 | 0.021 | Detected | Detected | 21.29 | True Positive |
| URV-130 | *Enterococcus faecalis* | 0.972 | 0.179 | Detected | Detected | 19.99 | True Positive |
| URV-132 | *Enterococcus faecalis* | 0.999 | 0.398 | Detected | Detected | 27.53 | True Positive |
| URV-133 | *Enterococcus faecalis* | 0.962 | 0.019 | Detected | Detected | 25.07 | True Positive |
| URV-134 | *Enterococcus faecalis* | 0.984 | 0.008 | Detected | Detected | 19.91 | True Positive |
| URV-137 | *Enterococcus faecalis* | 0.999 | 0.120 | Detected | Detected | 24.43 | True Positive |
| URV-139 | *Enterococcus faecalis* | 0.595 | 0.012 | Detected | Not_detected | Undetermined | False Positive |
| URV-146 | *Enterococcus faecalis* | 0.999 | 0.041 | Detected | Detected | 17.30 | True Positive |
| URV-157 | *Enterococcus faecalis* | 0.999 | 0.384 | Detected | Detected | 16.81 | True Positive |
| URV-163 | *Enterococcus faecalis* | 0.999 | 0.739 | Detected | Detected | 13.77 | True Positive |
| URV-170 | *Enterococcus faecalis* | 0.999 | 0.194 | Detected | Detected | 21.68 | True Positive |
| URV-172 | *Enterococcus faecalis* | 0.999 | 0.279 | Detected | Detected | 18.98 | True Positive |
| URV-176 | *Enterococcus faecalis* | 0.999 | 0.090 | Detected | Detected | 17.53 | True Positive |
| URV-180 | *Enterococcus faecalis* | 0.999 | 0.575 | Detected | Detected | 21.81 | True Positive |
| URV-181 | *Enterococcus faecalis* | 0.963 | 0.989 | Detected | Detected | 14.67 | True Positive |
| URV-182 | *Enterococcus faecalis* | 0.999 | 0.155 | Detected | Detected | 16.58 | True Positive |
| URV-184 | *Enterococcus faecalis* | 0.676 | 0.013 | Detected | Detected | 27.10 | True Positive |
| URV-190 | *Enterococcus faecalis* | 0.999 | 0.061 | Detected | Detected | 19.73 | True Positive |
| URV-220 | *Enterococcus faecalis* | 0.982 | 0.805 | Detected | Detected | 15.03 | True Positive |
| URV-226 | *Enterococcus faecalis* | 0.999 | 0.663 | Detected | Detected | 14.43 | True Positive |
| URV-227 | *Enterococcus faecalis* | 0.998 | 0.021 | Detected | Detected | 26.06 | True Positive |
| URV-232 | *Enterococcus faecalis* | 0.999 | 0.305 | Detected | Detected | 21.34 | True Positive |
| URV-248 | *Enterococcus faecalis* | 0.999 | 0.924 | Detected | Detected | 16.66 | True Positive |
| URV-258 | *Enterococcus faecalis* | 0.115 | 0.001 | Not detected | Detected | 24.59 | False Negative |
| URV-259 | *Enterococcus faecalis* | 0.402 | 0.013 | Not detected | Detected | 24.6 | False Negative |
| URV-261 | *Enterococcus faecalis* | 0.999 | 0.895 | Detected | Detected | 16.26 | True Positive |
| URV-272 | *Enterococcus faecalis* | 0.999 | 0.616 | Detected | Detected | 15.76 | True Positive |
| URV-278 | *Enterococcus faecalis* | 0.999 | 0.663 | Detected | Detected | 16.33 | True Positive |
| URV-280 | *Enterococcus faecalis* | 0.999 | 0.046 | Detected | Detected | 22.42 | True Positive |
| URV-283 | *Enterococcus faecalis* | 0.999 | 0.756 | Detected | Detected | 22.47 | True Positive |
| URV-288 | *Enterococcus faecalis* | 0.999 | 0.061 | Detected | Detected | 24.09 | True Positive |
| URV-291 | *Enterococcus faecalis* | 0.999 | 0.133 | Detected | Detected | 17.96 | True Positive |
| URV-111 | *Escherichia coli* | 0.999 | 0.115 | Detected | Detected | 15.96 | True Positive |
| URV-113 | *Escherichia coli* | 0.391 | 0.025 | Not detected | Detected | 19.96 | False Negative |
| URV-114 | *Escherichia coli* | 0.998 | 0.104 | Detected | Detected | 18.05 | True Positive |
| URV-117 | *Escherichia coli* | 0.999 | 0.020 | Detected | Detected | 18.10 | True Positive |
| URV-130 | *Escherichia coli* | 0.999 | 0.133 | Detected | Detected | 30.40 | True Positive |
| URV-131 | *Escherichia coli* | 0.999 | 0.840 | Detected | Detected | 17.06 | True Positive |
| URV-137 | *Escherichia coli* | 0.020 | 0.000 | Not detected | Not_detected | Undetermined | True Negative |
| URV-139 | *Escherichia coli* | 0.000 | 0.000 | Not detected | Not Detected | Undetermined | True Negative |
| URV-141 | *Escherichia coli* | 0.000 | 0.000 | Not detected | Not Detected | Undetermined | True Negative |
| URV-173 | *Escherichia coli* | 0.814 | 0.043 | Detected | Detected | 19.11 | True Positive |
| URV-178 | *Escherichia coli* | 0.971 | 0.098 | Detected | Detected | 18.50 | True Positive |
| URV-257 | *Escherichia coli* | 0.999 | 0.027 | Detected | Detected | 21.83 | True Positive |
| URV-258 | *Escherichia coli* | 0.999 | 0.713 | Detected | Detected | 11.46 | True Positive |
| URV-268 | *Escherichia coli* | 0.801 | 0.041 | Detected | Detected | 22.08 | True Positive |
| URV-280 | *Escherichia coli* | 0.997 | 0.675 | Detected | Detected | 19.01 | True Positive |
| URV-282 | *Escherichia coli* | 0.999 | 0.057 | Detected | Detected | 26.26 | True Positive |
| URV-289 | *Escherichia coli* | 0.999 | 0.677 | Detected | Detected | 14.22 | True Positive |

| | | | | | | | |
|---|---|---|---|---|---|---|---|
| URV-110 | *Klebsiella pneumoniae* | 0.801 | 0.012 | Detected | Detected | 15.81 | True Positive |
| URV-122 | *Klebsiella pneumoniae* | 0.999 | 0.159 | Detected | Detected | 14.59 | True Positive |
| URV-132 | *Klebsiella pneumoniae* | 0.046 | 0.000 | Detected | Detected | 37.81 | True Positive |
| URV-147 | *Klebsiella pneumoniae* | 0.999 | 0.071 | Detected | Detected | 26.79 | True Positive |
| URV-169 | *Klebsiella pneumoniae* | 0.953 | 0.043 | Detected | Detected | 14.82 | True Positive |
| URV-184 | *Klebsiella pneumoniae* | 0.999 | 0.140 | Detected | Detected | 21.88 | True Positive |
| URV-255 | *Klebsiella pneumoniae* | 0.781 | 0.002 | Detected | Detected | 25.35 | True Positive |
| URV-262 | *Klebsiella pneumoniae* | 0.955 | 0.003 | Detected | Detected | 23.69 | True Positive |
| URV-264 | *Klebsiella pneumoniae* | 0.422 | 0.004 | Not detected | Detected | 21.42 | False Negative |
| URV-280 | *Klebsiella pneumoniae* | 0.801 | 0.011 | Detected | Detected | 25.16 | True Positive |
| URV-289 | *Klebsiella pneumoniae* | 0.813 | 0.016 | Detected | Detected | 18.48 | True Positive |
| URV-187 | *Proteus mirabilis* | 0.895 | 0.008 | Detected | Not Detected | Undetermined | False Positive |
| URV-192 | *Proteus mirabilis* | 0.999 | 0.369 | Detected | Detected | 25.17 | True Positive |
| URV-228 | *Proteus mirabilis* | 0.999 | 0.069 | Detected | Detected | 21.74 | True Positive |
| URV-231 | *Proteus mirabilis* | 0.998 | 0.147 | Detected | Detected | 31.71 | True Positive |
| URV-268 | *Proteus mirabilis* | 0.998 | 0.075 | Detected | Detected | 34.07 | True Positive |
| URV-291 | *Staphylococcus aureus* | 0.978 | 0.109 | Detected | Not_detected | Undetermined | False Positive |
| URV-118 | *Aerococcus urinae* | 0.968 | 0.001 | Detected | Detected | 28.17 | True Positive |
| URV-130 | *Aerococcus urinae* | 0.662 | 0.001 | Detected | Detected | 23.52 | True Positive |
| URV-139 | *Aerococcus urinae* | 0.594 | 0.001 | Detected | Detected | 30.94 | True Positive |
| URV-187 | *Aerococcus urinae* | 0.986 | 0.005 | Detected | Detected | 26.56 | True Positive |
| URV-284 | *Aerococcus urinae* | 0.986 | 0.002 | Detected | Detected | 26.70 | True Positive |
| URV-285 | *Aerococcus urinae* | 0.802 | 0.001 | Detected | Detected | 23.11 | True Positive |
| URV-287 | *Aerococcus urinae* | 0.939 | 0.002 | Detected | Detected | 24.04 | True Positive |
| URV-291 | *Aerococcus urinae* | 0.967 | 0.001 | Detected | Detected | 19.91 | True Positive |
| URV-108 | *Gardnerella vaginalis* | 0.684 | 0.002 | Detected | Not Detected | Undetermined | False Positive |
| URV-114 | *Gardnerella vaginalis* | 0.930 | 0.028 | Detected | Detected | 18.83 | True Positive |
| URV-125 | *Gardnerella vaginalis* | 0.948 | 0.142 | Detected | Detected | 36.87 | True Positive |
| URV-128 | *Gardnerella vaginalis* | 0.696 | 0.029 | Detected | Not Detected | Undetermined | False Positive |
| URV-136 | *Gardnerella vaginalis* | 0.859 | 0.003 | Detected | Detected | 26.86 | True Positive |
| URV-140 | *Gardnerella vaginalis* | 0.934 | 0.084 | Detected | Detected | 22.10 | True Positive |
| URV-163 | *Gardnerella vaginalis* | 0.875 | 0.030 | Detected | Not Detected | Undetermined | False Positive |
| URV-167 | *Gardnerella vaginalis* | 0.964 | 0.004 | Detected | 23.927 | 23.927 | True Positive |
| URV-175 | *Gardnerella vaginalis* | 0.658 | 0.084 | Detected | Not Detected | Undetermined | False Positive |
| URV-181 | *Gardnerella vaginalis* | 0.803 | 0.002 | Detected | 21.28 | 21.28 | True Positive |
| URV-191 | *Gardnerella vaginalis* | 0.738 | 0.098 | Detected | 17.23 | 17.23 | True Positive |
| URV-223 | *Gardnerella vaginalis* | 0.970 | 0.013 | Detected | 31.97 | 31.97 | True Positive |
| URV-233 | *Gardnerella vaginalis* | 0.944 | 0.083 | Detected | 15.60 | 15.60 | True Positive |
| URV-256 | *Gardnerella vaginalis* | 0.840 | 0.545 | Detected | 20.08 | 20.08 | True Positive |
| URV-263 | *Gardnerella vaginalis* | 0.994 | 0.011 | Detected | 28.93 | 28.93 | True Positive |
| URV-278 | *Gardnerella vaginalis* | 0.638 | 0.031 | Detected | Not Detected | Undetermined | False Positive |
| URV-286 | *Gardnerella vaginalis* | 0.608 | 0.178 | Detected | Not Detected | Undetermined | False Positive |
| URV-104 | *Prevotella species* | 0.986 | 0.001 | Detected | Detected | 14.83 | True Positive |
| URV-112 | *Prevotella species* | 0.998 | 0.012 | Detected | Detected | 15.96 | True Positive |
| URV-163 | *Prevotella species* | 0.874 | 0.001 | Detected | Detected | 15.49 | True Positive |
| URV-249 | *Prevotella species* | 0.998 | 0.029 | Detected | Detected | 16.78 | True Positive |
| URV-259 | *Prevotella species* | 0.992 | 0.006 | Detected | Detected | 17.4 | True Positive |
| URV-270 | *Prevotella species* | 0.994 | 0.005 | Detected | Detected | 17.79 | True Positive |
| URV-271 | *Prevotella species* | 0.961 | 0.003 | Detected | Detected | 17.87 | True Positive |

**Sanger Sequencing Comparator Results**

| Sample | Analyte | Prediction Probability | Relative abundance | Biotia-ID Result | Comparator result | % Identity | Comparator Agreement |
|---|---|---|---|---|---|---|---|
| URV-146 | *Enterococcus faecium* | 0.998 | 0.017 | Detected | Detected | 100% | True Positive |
| URV-225 | *Enterococcus faecium* | 0.961 | 0.624 | Detected | Detected | 100% | True Positive |
| URV-176 | *Morganella morganii* | 0.988 | 0.004 | Detected | Detected | 100% | True Positive |
| URV-272 | *Morganella morganii* | 0.998 | 0.013 | Detected | Detected | 100% | True Positive |
| URV-112 | *aphylococcus epidermidi* | 0.828 | 0.001 | Detected | Detected | 100% | True Positive |
| URV-116 | *aphylococcus epidermidi* | 0.984 | 0.005 | Detected | Detected | 99.50% | True Positive |

| | | | | | | | |
|---|---|---|---|---|---|---|---|
| URV-291 | *phylococcus saprophytic* | 0.976 | 0.008 | Detected | Detected | 99.25% | True Positive |
| URV-146 | *Streptococcus agalactiae* | 0.999 | 0.122 | Detected | Detected | 100% | True Positive |
| URV-167 | *Streptococcus agalactiae* | 0.949 | 0.004 | Detected | Detected | 100% | True Positive |
| URV-186 | *Streptococcus agalactiae* | 0.930 | 0.003 | Detected | Detected | 100% | True Positive |
| URV-232 | *Streptococcus agalactiae* | 0.985 | 0.006 | Detected | Detected | 100% | True Positive |
| URV-291 | ginosus group Streptoco | 0.565 | 0.002 | Detected | Detected | 100% | True Positive |
| URV-140 | ginosus group Streptoco | 0.827 | 0.017 | Detected | Not Detected | NA | False Positive |
| URV-275 | ginosus group Streptoco | 0.515 | 0.002 | Detected | Not Detected | NA | False Positive |
| URV-287 | ginosus group Streptoco | 0.538 | 0.004 | Detected | Not Detected | NA | False Positive |
| URV-154 | *rynebacterium urealyticu* | 0.998 | 0.024 | Detected | Detected | 100% | True Positive |
| URV-227 | *rynebacterium urealyticu* | 0.963 | 0.002 | Detected | Detected | 100% | True Positive |
| URV-287 | *rynebacterium urealyticu* | 0.549 | 0.000 | Detected | Detected | 100% | True Positive |
| URV-288 | *rynebacterium urealyticu* | 0.645 | 0.001 | Detected | Detected | 100% | True Positive |
| URV-225 | *Candida albicans* | 0.980 | 0.042 | Detected | Detected | 100% | True Positive |
| URV-230 | *Candida albicans* | 0.980 | 0.314 | Detected | Detected | 100% | True Positive |
| URV-233 | *Candida albicans* | 0.986 | 0.087 | Detected | Detected | 99.8% | True Positive |
| URV-260 | *Candida albicans* | 0.943 | 0.019 | Detected | Detected | 100% | True Positive |
| URV-278 | *Candida albicans* | 0.996 | 0.081 | Detected | Detected | 97.50% | True Positive |
| SBU-103 | *Candida albicans* | 0.998 | 0.171 | Detected | Detected | 99.8% | True Positive |
| SBU-161 | *Candida albicans* | 0.598 | 0.017 | Detected | Detected | 99.8% | True Positive |
| SBU-176 | *Candida albicans* | 0.963 | 0.378 | Detected | Detected | 100.0% | True Positive |
| URV-148 | *Candida glabrata* | 0.982 | 0.679 | Detected | Detected | 99.9% | True Positive |
| URV-182 | *Candida glabrata* | 0.982 | 0.314 | Detected | Detected | 98.70% | True Positive |
| URV-262 | *Candida glabrata* | 0.982 | 0.661 | Detected | Detected | 99.9% | True Positive |
| URV-288 | *Candida glabrata* | 0.991 | 0.047 | Detected | Detected | 99.90% | True Positive |
| SBU-021 | *Candida glabrata* | 0.974 | 0.347 | Detected | Detected | 100.00% | True Positive |
| SBU-047 | *Candida glabrata* | 0.972 | 0.305 | Detected | Detected | 99.80% | True Positive |
| SBU-137 | *Candida glabrata* | 0.967 | 0.016 | Detected | Detected | 99.88% | True Positive |
| SBU-162 | *Candida glabrata* | 0.981 | 0.075 | Detected | Detected | 99.76% | True Positive |
| SBU-017 | *Candida tropicalis* | 0.733 | 0.108 | Detected | Detected | 99.80% | True Positive |
| SBU-018 | *Candida tropicalis* | 0.988 | 0.392 | Detected | Detected | 100.00% | True Positive |
| SBU-027 | *Candida tropicalis* | 0.977 | 0.269 | Detected | Detected | 99.80% | True Positive |
| SBU-074 | *Candida tropicalis* | 0.577 | 0.171 | Detected | Detected | 98.80% | True Positive |
| SBU-017 | *Candida krusei* | 0.696 | 0.058 | Detected | Detected | 100% | True Positive |

**Supplementary Table 6A.** Performance Characteristics fo Key Bacterial Urogenital Taxa Reported with BIOTIA-ID

| Key pathogen | Method | TP | TN | FN | FP | Total | Sensitivity | Specificity |
|---|---|---|---|---|---|---|---|---|
| *Escherichia coli* | Clinical | 39 | 204 | 1 | 1 | 245 | 97.50% | 99.51% |
| | Contrived | 157 | 1096 | 0 | 13 | 1266 | 100.00% | 98.83% |
| | *in silico* | 3 | 8239 | 0 | 24 | 8266 | 100.00% | 99.71% |
| | Total | 199 | 9539 | 1 | 38 | 9777 | 99.50% | 99.60% |
| *Klebsiella pneumoniae* | Clinical | 28 | 214 | 2 | 1 | 245 | 93.33% | 99.53% |
| | Contrived | 166 | 1099 | 0 | 1 | 1266 | 100.00% | 99.91% |
| | *in silico* | 3 | 8250 | 0 | 13 | 8266 | 100.00% | 99.84% |
| | Total | 197 | 9563 | 2 | 15 | 9777 | 98.99% | 99.84% |
| *Proteus mirabilis* | Clinical | 22 | 222 | 0 | 1 | 245 | 100.00% | 99.55% |
| | Contrived | 163 | 1103 | 0 | 0 | 1266 | 100.00% | 100.00% |
| | *in silico* | 3 | 8255 | 0 | 8 | 8266 | 100.00% | 99.90% |
| | Total | 188 | 9580 | 0 | 9 | 9777 | 100.00% | 99.91% |
| *Enterococcus faecalis* | Clinical | 61 | 181 | 2 | 1 | 245 | 96.83% | 99.45% |
| | Contrived | 147 | 1119 | 0 | 0 | 1266 | 100.00% | 100.00% |
| | *in silico* | 3 | 8263 | 0 | 0 | 8266 | 100.00% | 100.00% |
| | Total | 211 | 9563 | 2 | 1 | 9777 | 99.06% | 99.99% |
| *Staphylococcus aureus* | Clinical | 2 | 242 | 0 | 1 | 245 | 100.00% | 99.59% |
| | Contrived | 172 | 1094 | 0 | 0 | 1266 | 100.00% | 100.00% |
| | *in silico* | 3 | 8252 | 0 | 11 | 8266 | 100.00% | 99.87% |
| | Total | 177 | 9588 | 0 | 12 | 9777 | 100.00% | 99.88% |
| *Citrobacter* species | Clinical | 0 | 243 | 0 | 2 | 245 | - | 99.18% |
| | Contrived | 2 | 1264 | 0 | 0 | 1266 | 100.00% | 100.00% |
| | *in silico* | 20 | 8246 | 0 | 0 | 8266 | 100.0% | 100.0% |
| | Total | 22 | 9753 | 0 | 2 | 9777 | 100.00% | 99.98% |
| *Enterobacter aerogenes* | Clinical | 0 | 244 | 1 | 0 | 245 | 0.00% | 100.00% |
| | Contrived | 1 | 1265 | 0 | 0 | 1266 | 100.00% | 100.00% |
| | *in silico* | 3 | 8263 | 0 | 0 | 8266 | 100.00% | 100.00% |
| | Total | 4 | 9772 | 1 | 0 | 9777 | 80.00% | 100.00% |
| *Enterobacter cloacae* | Clinical | 0 | 245 | 0 | 0 | 245 | - | 100.00% |
| | Contrived | 1 | 1265 | 0 | 0 | 1266 | 100.00% | 100.00% |
| | *in silico* | 3 | 8253 | 0 | 10 | 8266 | 100.00% | 99.88% |
| | Total | 4 | 9763 | 0 | 10 | 9777 | 100.00% | 99.90% |
| *Klebsiella oxytoca* | Clinical | 0 | 244 | 0 | 1 | 245 | - | 99.59% |
| | Contrived | 1 | 1265 | 0 | 0 | 1266 | 100.00% | 100.00% |
| | *in silico* | 3 | 8260 | 0 | 3 | 8266 | 100.00% | 99.96% |
| | Total | 4 | 9769 | 0 | 4 | 9777 | 100.00% | 99.96% |
| *Klebsiella variicola* | Clinical | 1 | 242 | 0 | 2 | 245 | 100.00% | 99.18% |
| | Contrived | 1 | 1265 | 0 | 0 | 1266 | 100.00% | 100.00% |
| | *in silico* | 3 | 8260 | 0 | 3 | 8266 | 100.00% | 99.96% |
| | Total | 5 | 9767 | 0 | 5 | 9777 | 100.00% | 99.95% |
| *Morganella morganii* | Clinical | 2 | 243 | 0 | 0 | 245 | 100.00% | 100.00% |
| | Contrived | 2 | 1264 | 0 | 0 | 1266 | 100.00% | 100.00% |
| | in silico | 3 | 8262 | 0 | 1 | 8266 | 100.00% | 99.99% |
| | Total | 7 | 9769 | 0 | 1 | 9777 | 100.00% | 99.99% |
| *Proteus vulgaris* | Clinical | 0 | 244 | 1 | 0 | 245 | 0.00% | 100.00% |
| | Contrived | 1 | 1265 | 0 | 0 | 1266 | 100.00% | 100.00% |
| | *in silico* | 3 | 8256 | 0 | 7 | 8266 | 100.00% | 99.92% |
| | Total | 4 | 9765 | 1 | 7 | 9777 | 80.00% | 99.93% |
| *Providencia rettgeri* | Clinical | 0 | 244 | 0 | 1 | 245 | - | 99.59% |
| | Contrived | 1 | 1265 | 0 | 0 | 1266 | 100.00% | 100.00% |
| | *in silico* | 3 | 8260 | 0 | 3 | 8266 | 100.00% | 99.96% |
| | Total | 4 | 9769 | 0 | 4 | 9777 | 100.00% | 99.96% |
| *Providencia stuartii* | Clinical | 0 | 244 | 0 | 1 | 245 | - | 99.59% |
| | Contrived | 1 | 1265 | 0 | 0 | 1266 | 100.00% | 100.00% |
| | *in silico* | 3 | 8260 | 0 | 3 | 8266 | 100.00% | 99.96% |
| | Total | 4 | 9769 | 0 | 4 | 9777 | 100.00% | 99.96% |
| *Raoultella ornithinolytica* | Clinical | 0 | 245 | 0 | 0 | 245 | - | 100.00% |
| | Contrived | 1 | 1265 | 0 | 0 | 1266 | 100.00% | 100.00% |
| | *in silico* | 3 | 8260 | 0 | 3 | 8266 | 100.00% | 99.96% |
| | Total | 4 | 9770 | 0 | 3 | 9777 | 100.00% | 99.97% |
| *Serratia marcescens* | Clinical | 0 | 245 | 0 | 0 | 245 | - | 100.00% |
| | Contrived | 1 | 1265 | 0 | 0 | 1266 | 100.00% | 100.00% |
| | *in silico* | 3 | 8245 | 0 | 18 | 8266 | 100.00% | 99.78% |
| | Total | 4 | 9755 | 0 | 18 | 9777 | 100.00% | 99.82% |
| *Acinetobacter baumanii* | Clinical | 0 | 244 | 0 | 1 | 245 | - | 99.59% |
| | Contrived | 4 | 1262 | 0 | 0 | 1266 | 100.00% | 100.00% |
| | *in silico* | 3 | 8244 | 0 | 19 | 8266 | 100.00% | 99.77% |
| | Total | 7 | 9750 | 0 | 20 | 9777 | 100.00% | 99.80% |
| | Clinical | 0 | 245 | 0 | 0 | 245 | - | 100.00% |

| | | | | | | | | |
|---|---|---|---|---|---|---|---|---|
| *Acinetobacter lwolfii* | Contrived | 1 | 1265 | 0 | 0 | 1266 | 100.00% | 100.00% |
| | *in silico* | 3 | 8260 | 0 | 3 | 8266 | 100.0% | 100.0% |
| | Total | 4 | 9770 | 0 | 3 | 9777 | 100.00% | 99.97% |
| *Pseudomonas aeruginosa* | Clinical | 0 | 244 | 0 | 1 | 245 | - | 100% |
| | Contrived | 7 | 1259 | 0 | 0 | 1266 | 100.00% | 100.00% |
| | *in silico* | 3 | 8260 | 0 | 3 | 8266 | 100.0% | 100.0% |
| | Total | 10 | 9763 | 0 | 4 | 9777 | 100.00% | 99.96% |
| *Strenotrophomonas maltophilia* | Clinical | 0 | 244 | 0 | 1 | 245 | - | 100% |
| | Contrived | 1 | 1265 | 0 | 0 | 1266 | 100.00% | 100.00% |
| | *in silico* | 3 | 8252 | 0 | 11 | 8266 | 100.0% | 100.0% |
| | Total | 4 | 9761 | 0 | 12 | 9777 | 100.00% | 99.88% |
| *Aerococcus* species | Clinical | 8 | 237 | 0 | 0 | 245 | 100.00% | 100.00% |
| | Contrived | 2 | 1264 | 0 | 0 | 1266 | 100.00% | 100.00% |
| | *in silico* | 21 | 8242 | 3 | 0 | 8266 | 87.50% | 100.00% |
| | Total | 31 | 9743 | 3 | 0 | 9777 | 91.18% | 100.00% |
| Anginosus group Streptococci | Clinical | 1 | 235 | 0 | 9 | 245 | 100.00% | 96.31% |
| | Contrived | 1 | 1265 | 0 | 0 | 1266 | 100.00% | 100.00% |
| | *in silico* | 5 | 8261 | 0 | 0 | 8266 | 100.00% | 100.00% |
| | Total | 7 | 9761 | 0 | 9 | 9777 | 100.00% | 99.91% |
| *Corynebacterium urealyticum* | Clinical | 4 | 241 | 0 | 0 | 245 | 100.00% | 100.00% |
| | Contrived | 1 | 1265 | 0 | 0 | 1266 | 100.00% | 100.00% |
| | *in silico* | 3 | 8263 | 0 | 0 | 8266 | 100.00% | 100.00% |
| | Total | 8 | 9769 | 0 | 0 | 9777 | 100.00% | 100.00% |
| *Enterococcus faecium* | Clinical | 2 | 241 | 0 | 2 | 245 | 100.00% | 99.18% |
| | Contrived | 1 | 1003 | 0 | 0 | 1004 | 100.00% | 100.00% |
| | *in silico* | 3 | 8260 | 0 | 3 | 8266 | 100.00% | 99.96% |
| | Total | 6 | 9504 | 0 | 5 | 9515 | 100.00% | 99.95% |
| Mitis group Streptococci | Clinical | 0 | 245 | 0 | 0 | 245 | - | 100.00% |
| | Contrived | 1 | 1265 | 0 | 0 | 1266 | 100.00% | 100.00% |
| | *in silico* | 3 | 8262 | 0 | 1 | 8266 | 100.00% | 99.99% |
| | Total | 4 | 9772 | 0 | 1 | 9777 | 100.00% | 99.99% |
| *Staphylococcus epidermidis* | Clinical | 2 | 242 | 0 | 1 | 245 | 100.00% | 99.59% |
| | Contrived | 5 | 1261 | 0 | 0 | 1266 | 100.00% | 100.00% |
| | *in silico* | 3 | 8263 | 0 | 0 | 8266 | 100.00% | 100.00% |
| | Total | 10 | 9766 | 0 | 1 | 9777 | 100.00% | 99.99% |
| *Staphylococcus lugdunensis* | Clinical | 0 | 245 | 0 | 0 | 245 | - | 100.00% |
| | Contrived | 1 | 1265 | 0 | 0 | 1266 | 100.00% | 100.00% |
| | *in silico* | 3 | 8263 | 0 | 0 | 8266 | 100.00% | 100.00% |
| | Total | 4 | 9773 | 0 | 0 | 9777 | 100.00% | 100.00% |
| *Staphylococcus saprophyticus* | Clinical | 1 | 242 | 0 | 2 | 245 | 100.00% | 99.18% |
| | Contrived | 1 | 1265 | 0 | 0 | 1266 | 100.00% | 100.00% |
| | *in silico* | 3 | 8263 | 0 | 0 | 8266 | 100.00% | 100.00% |
| | Total | 5 | 9770 | 0 | 2 | 9777 | 100.00% | 99.98% |
| *Other Staphylococci* | Clinical | 0 | 245 | 0 | 0 | 245 | - | 100.00% |
| | Contrived | 9 | 1257 | 0 | 0 | 1266 | 100.00% | 100.00% |
| | *in silico* | 3 | 8263 | 0 | 0 | 8266 | 100.00% | 100.00% |
| | Total | 12 | 9765 | 0 | 0 | 9777 | 100.00% | 100.00% |
| *Streptococcus agalactiae* | Clinical | 4 | 241 | 0 | 0 | 245 | 100.00% | 100.00% |
| | Contrived | 1 | 1265 | 0 | 0 | 1266 | 100.00% | 100.00% |
| | *in silico* | 3 | 8263 | 0 | 0 | 8266 | 100.00% | 100.00% |
| | Total | 8 | 9769 | 0 | 0 | 9777 | 100.00% | 100.00% |
| *Anaerococcus vaginalis* | Clinical | 0 | 245 | 0 | 0 | 245 | - | 100.00% |
| | Contrived | 1 | 1265 | 0 | 0 | 1266 | 100.00% | 100.00% |
| | *in silico* | 3 | 8263 | 0 | 0 | 8266 | 100.00% | 100.00% |
| | Total | 4 | 9773 | 0 | 0 | 9777 | 100.00% | 100.00% |
| *Bacteroides fragilis* | Clinical | 0 | 245 | 0 | 0 | 245 | - | 100.00% |
| | Contrived | 7 | 1259 | 0 | 0 | 1266 | 100.00% | 100.00% |
| | *in silico* | 3 | 8263 | 0 | 0 | 8266 | 100.00% | 100.00% |
| | Total | 10 | 9767 | 0 | 0 | 9777 | 100.00% | 100.00% |
| *Prevotella* species | Clinical | 7 | 238 | 0 | 0 | 245 | 100.00% | 100.00% |
| | Contrived | 7 | 1259 | 0 | 0 | 1266 | 100.00% | 100.00% |
| | *in silico* | 9 | 8257 | 0 | 0 | 8266 | 100.00% | 100.00% |
| | Total | 23 | 9754 | 0 | 0 | 9777 | 100.00% | 100.00% |
| *Gardnerella vaginalis* | Clinical | 11 | 228 | 0 | 6 | 245 | 100.00% | 97.44% |
| | Contrived | 7 | 1259 | 0 | 0 | 1266 | 100.00% | 100.00% |
| | *in silico* | 3 | 8263 | 0 | 0 | 8266 | 100.00% | 100.00% |
| | Total | 21 | 9750 | 0 | 6 | 9777 | 100.00% | 99.94% |
| Negative Organisms | Clinical | 15 | 230 | 0 | 0 | 245 | 100.00% | 100.00% |
| | Contrived | 223 | 1230 | 0 | 0 | 1266 | 100.00% | 100.00% |
| | *in silico* | 7977 | 289 | 0 | 0 | 8266 | 100.00% | 100.00% |
| | Total | 8215 | 1749 | 0 | 0 | 9777 | 100.00% | 100.00% |

Supplementary Table 6B. Performance Characteristics fo Key Fungal Urogenital Taxa Reported with BIOTIA-ID

| Key pathogen | Method | TP | TN | FN | FP | Total | Sensitivity | Specificity |
|---|---|---|---|---|---|---|---|---|
| *Candida albicans* | Clinical | 8 | 28 | 0 | 0 | 36 | 100.0% | 100.0% |
| | Contrived | 132 | 795 | 1 | 0 | 928 | 99.2% | 100.0% |
| | *in silico* | 9 | 4133 | 0 | 0 | 4142 | 100.0% | 100.0% |
| | *Total* | 149 | 4956 | 1 | 0 | 5106 | 99.3% | 100.0% |
| *Candida auris* | Clinical | 0 | 36 | 0 | 0 | 36 | - | 100.0% |
| | Contrived | 129 | 797 | 0 | 2 | 928 | 100.0% | 99.7% |
| | *in silico* | 9 | 4133 | 0 | 0 | 4142 | 100.0% | 100.0% |
| | *Total* | 138 | 4966 | 0 | 2 | 5106 | 100.0% | 100.0% |
| *Candida dubliniensis* | Clinical | 0 | 36 | 0 | 0 | 36 | - | 100.0% |
| | Contrived | 10 | 914 | 0 | 0 | 924 | 100.0% | 100.0% |
| | *in silico* | 9 | 4133 | 0 | 0 | 4142 | 100.0% | 100.0% |
| | *Total* | 19 | 5083 | 0 | 0 | 5102 | 100.0% | 100.0% |
| *Candida glabrata* | Clinical | 8 | 28 | 0 | 0 | 36 | 100.0% | 100.0% |
| | Contrived | 135 | 793 | 0 | 0 | 928 | 100.0% | 100.0% |
| | *in silico* | 9 | 4133 | 0 | 0 | 4142 | 100.0% | 100.0% |
| | *Total* | 152 | 4954 | 0 | 0 | 5106 | 100.0% | 100.0% |
| *Candida guilliermondii* | Clinical | 0 | 36 | 0 | 0 | 36 | - | 100.0% |
| | Contrived | 10 | 918 | 0 | 0 | 928 | 100.0% | 100.0% |
| | *in silico* | 9 | 4133 | 0 | 0 | 4142 | 100.0% | 100.0% |
| | *Total* | 19 | 5087 | 0 | 0 | 5106 | 100.0% | 100.0% |
| *Candida kefyr* | Clinical | 0 | 36 | 0 | 0 | 36 | - | 100.0% |
| | Contrived | 10 | 918 | 0 | 0 | 928 | 100.0% | 100.0% |
| | *in silico* | 9 | 4133 | 0 | 0 | 4142 | 100.0% | 100.0% |
| | *Total* | 19 | 5087 | 0 | 0 | 5106 | 100.0% | 100.0% |
| *Candida krusei* | Clinical | 1 | 35 | 0 | 0 | 36 | - | 100.0% |
| | Contrived | 137 | 791 | 0 | 0 | 928 | 100.0% | 100.0% |
| | *in silico* | 9 | 4133 | 0 | 0 | 4142 | 100.0% | 100.0% |
| | *Total* | 147 | 4959 | 0 | 0 | 5106 | 100.0% | 100.0% |
| *Candida lusitaniae* | Clinical | 0 | 36 | 0 | 0 | 36 | - | 100.0% |
| | Contrived | 10 | 882 | 0 | 0 | 928 | 100.0% | 100.0% |
| | *in silico* | 9 | 4133 | 0 | 0 | 4142 | 100.0% | 100.0% |
| | *Total* | 19 | 5051 | 0 | 0 | 5106 | 100.0% | 100.0% |
| *Candida parapsilosis* | Clinical | 0 | 36 | 0 | 0 | 36 | - | 100.0% |
| | Contrived | 139 | 789 | 0 | 0 | 928 | 100.0% | 100.0% |
| | *in silico* | 9 | 4133 | 0 | 0 | 4142 | 100.0% | 100.0% |
| | *Total* | 148 | 4958 | 0 | 0 | 5106 | 100.0% | 100.0% |
| *Candida tropicalis* | Clinical | 4 | 32 | 0 | 0 | 36 | 100.0% | 100.0% |
| | Contrived | 134 | 794 | 0 | 0 | 928 | 100.0% | 100.0% |
| | *in silico* | 9 | 4133 | 0 | 0 | 4142 | 100.0% | 100.0% |
| | *Total* | 147 | 4959 | 0 | 0 | 5106 | 100.0% | 100.0% |
| Negative Organisms | Clinical | 9 | 191 | 0 | 0 | 200 | 100.0% | 100.0% |
| | Contrived | 79 | 849 | 0 | 0 | 928 | 100.0% | 100.0% |
| | *in silico* | 4052 | 90 | 0 | 0 | 4142 | 100.0% | 100.0% |
| | *Total* | 4140 | 1130 | 0 | 0 | 5270 | 100.0% | 100.0% |
